# Gut Microbiome and Risk of Dementia –a Prospective, Population-Based Study

**DOI:** 10.64898/2026.02.15.26345196

**Authors:** J. Tynkkynen, O. Kambur, T. Niiranen, L. Lahti, M. Ruuskanen, D. McDonald, P. Jousilahti, C. Gazolla Volpiano, G. Méric, M. Inouye, Y. Liu, L. Khatib, L. Patel, V. Salomaa, R. Knight, A.S. Havulinna

## Abstract

**INTRODUCTION:** The pathophysiology and risk factors for Alzheimer’s disease (AD) and dementia are insufficiently known. We studied the connections between gut microbiome, overall dementia and AD in a prospective, population-based cohort.

**METHODS:** We followed a population based random sample of 4,055 individuals (FINRISK 2022) for 16 years, with 330 cases of incident dementia and 280 AD cases. Gut microbiome community diversity and composition were assessed against future dementia and AD risk. Competing mortality risks were accounted for using Fine–Gray models.

**RESULTS:** Community diversity was not associated with dementia or AD. However, a supervised ordination with dbRDA suggested a possible compositional link between gut microbiome and dementia. One putative bacterial genus, *Dorea*, was associated with a decreased dementia risk. APOE ε4 genotype associated with several taxa; of these, phylum *Verrucomicrobiota* and species *Nocardia carnea* were associated with incident dementia.

**DISCUSSION:** The gut-brain axis has a modest association on future dementia or AD risk. Microbial composition, rather diversities, may contribute to dementia risk.

## INTRODUCTION

Alzheimer’s disease (AD) is the most common cause of dementia. The pathological process of AD is considered to have multiple interplays between genetic and environmental risk factors (1) and infectious diseases may affect the AD process. In post-mortem studies, several viral, bacterial, and even fungal residues have been found directly in brain samples of AD patients(2). Increased levels of amyloid beta (Aβ) accumulation are detected in herpes simplex type 1 infected mice as well as in cultured human brain cells, indicating a lifelong infection burden to possibly initiate the AD process (3,4). Interestingly, the association between herpes virus infection and AD seems to be modified by the APOE ε4 genotype (5). Additionally, a slow spirochete infection has been suggested to be linked to AD pathogenesis (6).

Indirect microbial mechanisms may also affect brain functioning, and the gut microbiota alterations are associated with many neurological disorders(7). Cross-sectional studies show varying within-community diversity (α-) or composition (β-diversity) of gut microbiome among patients with AD, mild cognitive impairment (MCI), and healthy controls(8,9). Microbial α- and β-diversity, pro-inflammatory cytokines, and increased relative abundance of *Escherichia* and *Shigella* bacteria are connected with brain Aβ accumulation (10). Moreover, gut microbiome related amyloid proteins detected in fecal samples may trigger or accelerate the neurodegenerative process (11). Of individual taxa, the abundance of the *Enterobacteriaceae* family discriminated AD patients from controls (9). Several phylum-, family-, and genus-level differences in gut microbiome abundances have been observed between patients with AD and healthy controls, but these findings are inconsistent across studies (8–10,12–16).

Nearly all existing human studies are cross-sectional, limiting causal inference and raising the possibility that observed microbiome differences reflect disease consequences rather than antecedent risk factors. In our previous analysis focusing on human polygenic risk scores, the microbiome score did not improve AD risk discrimination during the follow-up of 16 years (17). Using the population-based FINRISK 2002 cohort with 16-year follow-up, we now study possible connections between gut microbiomes, APOE genotype, and future incidence of AD and dementia, in more detail.

## METHODS

### Study population and outcome

The FINRISK 2002 is a population-based health examination survey conducted by The Finnish Institute for Health and Welfare (18). A random population sample of 13,437 persons from six geographic regions, aged between 25 and 74 years, was invited, and 8,738 subjects participated in the survey. All participants completed a questionnaire on demographic information, lifestyles, medications, and medical history. Health examination was performed on all participants, and blood samples were taken at the local study sites. Stool samples were collected at home and sent to the Finnish Institute for Health and Welfare using prepaid postal packages over 1 to 2 days under typical Finnish winter conditions (19). Incident and prevalent diagnoses, including AD and other dementia diagnoses and deaths, were detected based on nation-wide Finnish health care registers: the Care Register for Health Care, the Causes of Death Register, and the National Social Insurance Institution’s Drug Reimbursement Register (20). The overall accuracy of these registers is considered good (21), and this methodology provides precise detection of AD and dementia cases but lacks some sensitivity (22). Finnish electronic health care (EH) - registers cover 100% of all prescribed drug purchases, drug reimbursements, hospitalisations, and deaths in Finland. ICD-10 codes F00 (AD related dementia), F01 (vascular dementia), F02 (dementia in other diseases), F03 (unspecified dementia), and G30 (AD) were used to detect dementia. Thus, our diagnosis of incident overall dementia included AD (amyloidopathy) and vascular dementia but excluded Parkinson’s dementia, the most common alpha-synucleinpathy. Incident AD was identified if the drug reimbursement was granted based on a statement written by a specialist physician (neurologist or geriatrician) and approved by the National Social Insurance Institution. AD diagnoses are based on the Finnish Current Care Guidelines (23) recommendation, which follow the NINCDS-ADRDA criteria (24).

The follow-up continued until December 31^st^, 2018, or until AD, dementia or death was detected. We analysed the results only among study subjects over 40 years of age at the time of stool collection to focus on the population at actual risk for incident AD or dementia. Study subjects who did not provide a stool sample (n=1,507) or had a low read count in their stool sample (n=20) were excluded from the analysis. Also, subjects who had been previously diagnosed with AD, dementia or Parkinson’s disease (n=36) or were pregnant during the survey (n=39) were excluded from the analysis. Subjects with non-recurring dementia diagnoses recorded only in the casualty department were excluded from the analysis (n=36) since diagnostics was not based on sufficient neurological work-up. The total sample size for this study was 4,055 subjects with known APOE ε4 genotype. The FINRISK 2002 study was approved by the ethics committee of Helsinki and Uusimaa Hospital District (decision number 87/2001), and all the study participants gave a written informed consent. The study was performed according to the Declaration of Helsinki.

### Fecal metagenomics

The collection and analysis of stool samples from the FINRISK 2002 cohort have been described previously (18). Briefly, stool samples were stored at -20°C prior to metagenomic sequencing in 2017 at the University of California, San Diego. Shallow shotgun metagenomic sequencing was performed, and subsequent FASTQ files were host-filtered and mapped against the Web of Life release 2 database using the SHOGUN v1.0.5 parameters, followed by filtering against Greengenes2 v2022.10 (25–27).

We investigated the variation in microbial community composition between samples with compositional data analysis (CoDA) methods using the Aitchison distance. To achieve this, we transformed the abundance count data using the centered log-ratio (CLR) method and then calculated the Euclidean distance between the CLR-transformed samples (28,29). CLR transform applies a pseudocount equal to half of the minimum non-zero relative abudance to zero entries in the OTU table to allow log-transformation. R/Bioconductor package microbiome (version 1.8.0) was used to calculate community diversity (Shannon index) and for transformations of microbial abundances (30). R package vegan (version 2.5.7) was used to calculate Aitchison distance (31). Principal component analysis (PCA) with the first three components was used in community composition and microbial PC analyses. PCA was performed using the R-package stats (version 3.6.2) (32). For α-diversity and microbial PC analyses, species-level data was used. Distance-based redundancy analysis (dbRDA) was calculated with vegan package using Aitchison distance matrix.

### Statistical analysis

The associations between gut microbiomes and incident AD and dementia were estimated with the Fine-Gray (FG) competing risks model. Death from other causes was considered a competing risk for AD and dementia. The FG model was adjusted for sex, the number of APOE ε4 alleles (0-2), self-reported duration of education in years graded into tertiles and adjusted for birth year (33), body mass index (BMI, kg/m^2^), mean systolic blood pressure of two measurements (mmHg), alcohol consumption (g/week), smoking status (current, ex-smoker, never), usage of anti-hypertensive medication, prevalent diabetes and cardiovascular disease (coronary heart disease, stroke, heart failure, atrial fibrillation or peripheral artery disease) and use of potentially gut microbiome altering drugs (medicine purchase including antibiotics of ATC-classes A, B, C, G, H, J, L, N and P within 0-4 months prior to the study baseline, Supplementary Table 1). Participant’s age was used as the time scale. The stool samples were collected and adjusting covariates recorded at baseline during the winter and early spring of 2002.

The significance level of α = 0.05 was used. We also tested the associations between all taxa detected in >1% of subjects (non-zero relative abundance in >1% of samples) and AD or dementia. P-values obtained from this analysis, including phylum, order, family, genus, and species, were corrected with false discovery rate analysis (FDR, q-value) using the Benjamini & Hochberg method.

The association between gut microbiomes and APOE genotype was studied with a linear regression model adjusted for age, sex, education level (described above), BMI, systolic blood pressure, alcohol consumption (g/week), smoking status (current, ex-smoker, never), usage of anti-hypertensive medication, prevalent diabetes, cardiovascular disease (described above) and use of potentially gut microbiome altering drugs (Supplementary Table 1). All taxa associated with APOE genotype (p<0.05) were further tested against AD or dementia risk and adjusted as described above, except APOE genotype was used as a binary variable (0 vs 1-2 ε4 alleles).

To test the microbiome composition between participants with and with-out incident dementia or AD, we performed dbRDA with PERMANOVA with 999 permutations. Models were adjusted for age, sex, number of APOE ε4 alleles, education level, BMI, systolic blood pressure, alcohol consumption, smoking status, usage of anti-hypertensive medication, gut microbiome altering drugs, prevalent diabetes and cardiovascular disease (CVD).

Extreme gradient boosting models were implemented using the XGBoost R package (v0.90.0.2) (34). Data were randomly split into training (70%) and validation (30%) sets. Model development, including hyperparameter tuning via cross-validation, was performed exclusively within the training set to prevent data leakage. Hyperparameters for XGBoost models were tuned and optimized inside the training set using the GridSearchCV package in scikit-learn. Final models were evaluated in the independent validation set. The outcome was Alzheimer’s disease/dementia case status (binary). Covariates matched those used in conventional regression analyses, and models were fitted with and without microbiome features. Microbial relative abundances were centered log-ratio (CLR) transformed prior to analysis. Model performance was assessed using the binary classification and Area Under the Curve (AUC) for the dementia risk prediction.

## RESULTS

During the median 15.8 years of follow-up, we detected 330 incident cases of dementia, of which 280 met the AD criteria. The baseline characteristics of the study participants are presented in Table 1. As expected, subjects with incident AD and dementia were older and were more likely APOE ε4 heterozygotes (intermediate risk) or ε4/ε4 homozygotes (high risk) than healthy controls. Incident AD and dementia cases had higher baseline BMI, systolic blood pressure, used anti-hypertensive and gut microbiome altering medication more frequently, and had diabetes or CVD more often. These associations were no longer observed in the FG regression models (Supplementary Table 2). Current smoking did not associate with the risk of incident AD or dementia in the FG analysis, but women were at lower dementia risk (Supplementary Table 2).

### Community diversity and Aitchison distance community composition

Community diversity (Shannon index) of the gut microbiome was not associated with incident AD or dementia risk. The hazard ratios for AD and dementia per one SD change in the Shannon α-diversity index were 1.10 (95% CI 0.98-1.23, p=0.105) and 1.07 (95% CI 0.96-1.20, p=0.225).

Future dementia development was associated with variation in microbiome composition based on Aitchison distance dbRDA in phylum, genus and species levels (Supplementary Table 3). In contrast, Aitchison distance PCA did not reveal any significant associations (Supplementary Table 4). Future development of AD, on the other hand, did not affect dbRDA. As a sensitivity analysis, we tested community diversity and composition in a model restricted to a ten-year follow-up and adjusted for binary APOE status (0 vs 1-2 ε4 alleles), but no statistically significant associations were observed (data not shown).

Additionally, none of the first three principal components of taxonomic abundance was associated with incident AD or dementia (Supplementary Table 5).

### Putative microbes based on previous literature

We tested, if putative microbes reported in the previous literature showed associations with incident AD or dementia. At the phylum or family level, no associations were observed. At the genera level, genus *Dorea* abundance was associated with lower dementia risk (HR 0.90, 95% CI 0.81-0.99, p=0.038) but not with AD risk (HR 0.92, 95% CI 0.83-1.03, p=0.167) (Supplementary Table 6). We also tested the associations between all taxa detected in >1% of subjects and AD or dementia. Type I error was controlled by FDR correction of the p-values. The genus *Dietzia* (HR 1.17, 95%CI 1.09-1.25, p<0.001, q-value=0.013) and species *Brevibacterium epidermidis* (HR 1.08, 95%CI 1.05-1.11, p<0.001, q-value=0.001) and *Liquorilactobacillus vini* (HR 1.17, 95%CI 1.09-1.26, p<0.001, q-value=0.019) were associated with dementia (Table 2). While the HRs for AD were similar for *Brevibacterium epidermidis* and *Liquorilactobacillus vini*, these associations did not remain significant after FDR correction.

The sensitivity analysis, with ten years follow-up time restriction and the model adjusted for binary APOE status, showed a *Brevibacterium epidermidis* association with increased dementia risk among with *Corynebacterium stationis, Leucobacter, Dietzia cinnamea, Corynebacterium efficiens, Mobilicoccus pelagius, Acinetobacter bouvetii* species (Supplementary Table 7), but not with AD risk

### Microbes associated with the APOE genotype and with incident dementia

Next, we tested if the APOE genotype – the most significant genetic risk factor for AD – would associate with gut microbiome and if these microbes would associate with AD or dementia risk. These latter analyses underwent FDR correction and were adjusted for APOE genotype as a binary variable (0 vs 1-2 ε4 alleles). In the linear regression model, the APOE genotype (number of ε4 alleles) was associated with several bacterial relative abundances at all taxonomic levels tested (Supplementary Table 8). Of these taxa, the phylum *Verrucomicrobiota* was associated with an increased risk of dementia (HR 1.20, 95% CI 1.03-1.39, q-value=0.040), and the results regarding AD were fairly similar, even though the associations were not statistically significant after FDR correction (HR 1.19, 95% CI 1.02-1.39, q-value=0.054) (Table 3). Additionally, this association seemed to extend to subordinate taxa as well: order *Verrucomicrobiales* (HR 1.22, 95% CI 1.05-1.41, p=0.010, q-value=0.060) (Table 3), family *Akkermansiaceae* (HR 1.22, 95%CI 1.05-1.41, p=0.010, q-value=0.190) (Supplementary Table 9), genus *Akkermansia* (HR 1.21, 95%CI 1.05-1.41, p=0.010, q-value=0.228) (Supplement Table 10) and species *Akkermansia glycaniphila* (HR 1.21, 95%CI 1.05-1.40, p=0.009, q-value=0.315) (Supplementary Table 11) were all nominally associated with increased AD risk, but not when the results were FDR corrected. Species *Nocardia carnea* was associated with dementia risk after FDR correction (HR 1.13, 95%CI 1.07-1.19, p<0.001, q-value<0.001).

### Machine learning models

Lastly, we trained machine learning prediction models (XGBoost) using taxonomic features at the phylum, genus, and species levels to predict future AD and dementia risk. However, the taxonomic profiles did not improve model performance over traditional risk factors such as age, APOE genotype, or sex (Supplementary Table 12). The phylum contributing most to the XGBoost model was *Bacteroidota*, yet ranked below age, APOE genotype, alcohol consumption, systolic blood pressure, and the use of gut microbiome-altering drugs (Supplementary Table 13). At the genus and species levels, *Granulicoccus* and *Prevotella oulorum* were the most contributing factors.

## DISCUSSION

This large population-based study investigated the associations between gut microbiomes and risk of AD and dementia in a prospective setting in a cohort of 4,055 participants and 280 incident AD and 330 dementia cases during 16 years of follow-up. Although not all-cross sectional studies have linked α- and β-diversities to amyloid–β pathogenesis (35), several small cross-sectional studies have demonstrated differences in α- and β-diversities between AD or MCI patients, and healthy controls (8–10,35). However, in our earlier analyses, we discovered no evident connection between AD risk and gut microbiome diversities or scores (17). The present, more focused and more in-depth investigation extended and confirmed those findings. On the other hand, dbRDA with Aitchison distance showed that future dementia, but not AD, associates with gut microbiome community composition though we were unable to show the association in time-to-event analysis with principal components. Compared to our previous study, we now used the Fine-Gray subdistribution model to cover the competing risk of deaths not related to AD or dementia and all our results were adjusted for APOE genotype and several clinical AD and dementia risk factors.

Proteomic studies have recently reported convincing associations between the APOE ε4 allele, inflammation, and a wide range of neurodegenerative diseases, including AD (36). We identified several microbial taxa associated with the APOE genotype. Of these taxa, phylum *Verrucomicrobiota* and its subphylum taxa were related to an increased risk of AD and dementia. Lower abundance of *Akkermansia muciniphila* has been previously linked to diabetes (37), obesity, hyperlipidemia (38), and fatty liver disease (39) and a higher abundance of *Akkermansia muciniphila* often seems to protect against these metabolic disorders (40–42). Additionally, cross-sectional human studies (43) and animal experiments show that *Akkermansia* could positively impact neuronal health (44,45). *Akkermansia muciniphila* is observed to reduce amyloid β accumulation in the cerebral cortex (46) and to improve cognitive functions in AD animal models. Yet, opposite results have also been published (47,48), and connections between *Verrucomicrobiota*, its subphylum taxa, and neurodegenerative disorders remain unclear. It should also be acknowledged that taxa at species, genus, family, and phyla exhibit substantial functional heterogeneity, which may partly explain the inconsistencies between previous studies and the present findings (47), as we did not observe any associations between *Akkermansia muciniphila* and the risk of dementia or AD.

Diet affects the gut microbiome status and the abundance of *Akkermansia* (45). Although not intentional, possible dietary changes in the subclinical phase of the neurodegenerative disease can impact the patient’s microbiome status. Unfortunately, this is a matter that we could not control for in this prospective follow-up study based on EHR endpoint detection. Nevertheless, this speculative effect on the results is smaller than in previous cross-sectional studies since we followed a random population sample without prevalent AD or dementia diagnoses at baseline.

Analyses based on putative microbes showed an association between the genus *Dorea* and decreased dementia risk in the current study. Previously, this association was observed to be the opposite (12), but a more recent small cross-sectional study demonstrated a similar association to ours (50). Moreover, species *Dorea longicatena* seems to have a positive effect on muscle mass and bone mineral density (51). On the other hand, genus *Dorea* has been linked to metabolic disorders like higher serum fasting glucose (52) and fatty liver disease (53), which cannot be considered protective factors for AD or dementia. However, in our analyses, we accounted for various clinical factors, such as prevalent diabetes and BMI, thereby addressing the potential confounding effects of metabolic disorders on our results. Although correlations with dementia were detected, the statistical significance was not observed for AD. This is likely due to the slightly lower number of AD cases compared to overall dementia cases (280 vs. 330), given that the point estimates were comparable between dementia and AD analyses. Opposite to genus *Dorea*, no other putative microbes were associated with incident AD or dementia. All previous studies are cross-sectional and have markedly smaller sample sizes, which may yield varying results compared to current study. On the other hand, we must consider contemporary AD and future AD not to involve the same biomarkers, while it seems that antibiotic consumption, which most likely affects gut microbiome, is not associated with increased dementia or cognitive decline risk (54). Finally, the human microbiota changes with age and becomes less stable in response to effects caused by, for example, medication use and dietary changes (55,56).

When we analyzed all taxa detected >1% in the study population, only the genus *Dietzia* and species *Brevibacterium epidermidis* and *Liquorilactobacillus vini* were associated with increased dementia, but not AD risk. Genus *Dietzia* is an opportunistic human pathogen causing skin and soft tissue infections (57,58) but no previous associations with dementia have been reported. Neither *Brevibacterium epidermidis* nor *Liquorilactobacillus vini* has been connected to neurodegenerative disorders or reported to have significant pathogenic properties. Therefore, these observations should be interpreted with caution. Species *Nocardia carnea*, linked to APOE genotype and increased dementia risk in this study, causes mainly fulminant infections in immunosuppressed patients (59), and similar caution should be exercised in interpreting these findings.

This study has several strengths. First, we employed 16 years of follow-up with a randomly selected, representative population sample. The coverage of follow-up is almost 100% in this population cohort. This, along with a study cohort of 4,055 participants, results in a unique opportunity to investigate associations between gut microbiome and neurodegenerative diseases. We adjusted our analyses for several clinical risk factors and APOE genotype. We also used the Fine-Gray competing risk model to control for the competing risk of death, which is relevant in the population at risk of dementia. Machine learning models were used to discover more complex connections between gut microbiome and dementia risk, but none of the taxa seemed to improve model performance over known risk factors. Lastly, our microbiome data were generated using untargeted shallow shotgun metagenomic sequencing, which generally provides higher accuracy than 16S-based approach, though with some considerations (60,61). Taxonomic mapping was primarily performed using Greengenes2 2022.10 reference tree.

This study also has limitations, which should be considered. First, AD and dementia detection relied on Finnish national EHRs, causing some lack of sensitivity and possible delay in the endpoint recording and detection. Secondly, it is necessary to replicate these results in an independent cohort to validate the findings from the large-scale data analysis. However, population-specific differences can influence gut microbiome, and microbiome-disease associations, potentially affecting replication of observations (62,63). Importantly, additional study cohorts could give more statistical power to uncover additive associations between the gut microbiome and AD risk e.g., in APOE ε4 subgroups and a simple presence–absence microbiome data transformation could also reveal some new associations between gut microbiome and dementia risk (64). On the other hand, we believe that our study was sufficiently powered to detect clinically relevant associations between gut microbiome diversities or abundances and AD and dementia risk. Unlike Fine-Gray models, community based ML or dbRDA models were not based on time-to-event analyses. On the other hand, it must be recognised that dementia and AD are slowly developing diseases for which the exact date of onset cannot be given. We have date of diagnosis in our database but the time interval between the onset of symptoms and the final diagnosis of dementia or AD can vary considerably, depending on circumstances.

Lastly, our analyses did not include viruses or fungi, which are also hypothesized to play a role in the development of AD and dementia (3,4,65). Only simple associations between gut microbiome and AD or dementia risk were examined. Testing interactions between taxa would rapidly increase complexity of the models. The interplay between gut microbiome and circulating metabolites should also be studied in the future.

In conclusion, the connection between the gut microbiome and AD or dementia risk appears to be limited when considering community diversity or composition of gut microbiome abundance. This study, along with previous cross-sectional research, suggests that while the gut microbiome may not affect dementia risk greatly, dementia and MCI might affect the gut microbiome. On the other hand, some observations were made in this prospective setting: the ε4 genotype is associated with several microbial taxa, some of which may underlie the compositional findings observed. *Verrucomicrobiota* and its subordinate taxa were associated with higher AD and dementia risk, even though the magnitude of the risk was modest compared to more established risk factors. This observation is opposite to most findings in previous cross-sectional studies, and like all ‘omics’ analyses, all these observations warrants careful experimental validation.

## Data Availability

All data produced in the present study are available upon reasonable request to the authors

## References

1. Lane CA, Hardy J, Schott JM. Alzheimer’s disease. Eur J Neurol. 2018 Jan;25(1):59–70.

2. Salihoğlu R, Önal-Süzek T. Tissue Microbiome Associated With Human Diseases by Whole Transcriptome Sequencing and 16S Metagenomics. Front Genet. 2021;12:585556.

3. Wozniak MA, Itzhaki RF, Shipley SJ, Dobson CB. Herpes simplex virus infection causes cellular beta-amyloid accumulation and secretase upregulation. Neurosci Lett. 2007 Dec 18;429(2–3):95–100.

4. Eimer WA, Vijaya Kumar DK, Navalpur Shanmugam NK, Rodriguez AS, Mitchell T, Washicosky KJ, et al. Alzheimer’s Disease-Associated β-Amyloid Is Rapidly Seeded by Herpesviridae to Protect against Brain Infection. Neuron. 2018 Jul 11;99(1):56-63.e3.

5. Linard M, Letenneur L, Garrigue I, Doize A, Dartigues JF, Helmer C. Interaction between APOE4 and herpes simplex virus type 1 in Alzheimer’s disease. Alzheimers Dement J Alzheimers Assoc. 2020 Jan;16(1):200–8.

6. Miklossy J. Alzheimer’s disease - a neurospirochetosis. Analysis of the evidence following Koch’s and Hill’s criteria. J Neuroinflammation. 2011 Aug 4;8:90.

7. Cryan JF, O’Riordan KJ, Sandhu K, Peterson V, Dinan TG. The gut microbiome in neurological disorders. Lancet Neurol. 2020 Feb;19(2):179–94.

8. Vogt NM, Kerby RL, Dill-McFarland KA, Harding SJ, Merluzzi AP, Johnson SC, et al. Gut microbiome alterations in Alzheimer’s disease. Sci Rep. 2017 Oct 19;7(1):13537.

9. Liu P, Wu L, Peng G, Han Y, Tang R, Ge J, et al. Altered microbiomes distinguish Alzheimer’s disease from amnestic mild cognitive impairment and health in a Chinese cohort. Brain Behav Immun. 2019 Aug;80:633–43.

10. Cattaneo A, Cattane N, Galluzzi S, Provasi S, Lopizzo N, Festari C, et al. Association of brain amyloidosis with pro-inflammatory gut bacterial taxa and peripheral inflammation markers in cognitively impaired elderly. Neurobiol Aging. 2017 Jan;49:60–8.

11. Fernández-Calvet A, Matilla-Cuenca L, Izco M, Navarro S, Serrano M, Ventura S, et al. Gut microbiota produces biofilm-associated amyloids with potential for neurodegeneration. Nat Commun. 2024 May 16;15(1):4150.

12. Li B, He Y, Ma J, Huang P, Du J, Cao L, et al. Mild cognitive impairment has similar alterations as Alzheimer’s disease in gut microbiota. Alzheimers Dement J Alzheimers Assoc. 2019 Oct;15(10):1357–66.

13. Saji N, Niida S, Murotani K, Hisada T, Tsuduki T, Sugimoto T, et al. Analysis of the relationship between the gut microbiome and dementia: a cross-sectional study conducted in Japan. Sci Rep. 2019 Jan 30;9(1):1008.

14. Saji N, Murotani K, Hisada T, Tsuduki T, Sugimoto T, Kimura A, et al. The Association between Cerebral Small Vessel Disease and the Gut Microbiome: A Cross-Sectional Analysis. J Stroke Cerebrovasc Dis Off J Natl Stroke Assoc. 2021 Mar;30(3):105568.

15. Ning J, Huang SY, Chen SD, Zhang YR, Huang YY, Yu JT. Investigating Casual Associations Among Gut Microbiota, Metabolites, and Neurodegenerative Diseases: A Mendelian Randomization Study. J Alzheimers Dis JAD. 2022;87(1):211–22.

16. Cammann D, Lu Y, Cummings MJ, Zhang ML, Cue JM, Do J, et al. Genetic correlations between Alzheimer’s disease and gut microbiome genera. Sci Rep. 2023 Mar 31;13:5258.

17. Liu Y, Ritchie SC, Teo SM, Ruuskanen MO, Kambur O, Zhu Q, et al. Integration of polygenic and gut metagenomic risk prediction for common diseases. Nat Aging. 2024 Apr;4(4):584–94.

18. Borodulin K, Tolonen H, Jousilahti P, Jula A, Juolevi A, Koskinen S, et al. Cohort Profile: The National FINRISK Study. Int J Epidemiol. 2018 Jun 1;47(3):696–696i.

19. Palmu J, Salosensaari A, Havulinna AS, Cheng S, Inouye M, Jain M, et al. Association Between the Gut Microbiota and Blood Pressure in a Population Cohort of 6953 Individuals. J Am Heart Assoc. 2020 Aug 4;9(15):e016641.

20. Tynkkynen J, Laatikainen T, Salomaa V, Havulinna AS, Blankenberg S, Zeller T, et al. NT-proBNP and the risk of dementia: a prospective cohort study with 14 years of follow-up. J Alzheimers Dis JAD. 2015;44(3):1007–13.

21. Sund R. Quality of the Finnish Hospital Discharge Register: A systematic review. Scand J Public Health. 2012 Aug 1;40(6):505–15.

22. Solomon A, Ngandu T, Soininen H, Hallikainen MM, Kivipelto M, Laatikainen T. Validity of dementia and Alzheimer’s disease diagnoses in Finnish national registers. Alzheimers Dement J Alzheimers Assoc. 2014 May;10(3):303–9.

23. Working group set up by the Finnish Medical Society Duodecim, the Finnish geriatric association, the Finnish neurological association, the Finnish psychiatric association and the Finnish general medicine association. Memory disorders and dementia. Current Care Guidelines. Helsinki Finnis Medical society Duodecim 2020.

24. Dubois B, Feldman HH, Jacova C, Dekosky ST, Barberger-Gateau P, Cummings J, et al. Research criteria for the diagnosis of Alzheimer’s disease: revising the NINCDS-ADRDA criteria. The LancetNeurology. 2007 elokuu;6(8):734–46.

25. McDonald D, Jiang Y, Balaban M, Cantrell K, Zhu Q, Gonzalez A, et al. Greengenes2 unifies microbial data in a single reference tree. Nat Biotechnol. 2024 May;42(5):715–8.

26. Hillmann B, Al-Ghalith GA, Shields-Cutler RR, Zhu Q, Gohl DM, Beckman KB, et al. Evaluating the Information Content of Shallow Shotgun Metagenomics. mSystems. 2018 Dec;3(6).

27. O’Leary NA, Wright MW, Brister JR, Ciufo S, Haddad D, McVeigh R, et al. Reference sequence (RefSeq) database at NCBI: current status, taxonomic expansion, and functional annotation. Nucleic Acids Res. 2016 Jan 4;44(D1):D733–745.

28. Gloor GB, Macklaim JM, Pawlowsky-Glahn V, Egozcue JJ. Microbiome Datasets Are Compositional: And This Is Not Optional. Front Microbiol. 2017;8:2224.

29. Chen B, He X, Pan B, Zou X, You N. Comparison of beta diversity measures in clustering the high-dimensional microbial data. PloS One. 2021;16(2):e0246893.

30. Lahti L, Sudarshan S. Tools for microbiome analysis in R. Version. 2017. Available from: http://microbiome.github.com/microbiome.

31. Oksanen J, Blanchet FG, Friendly M, Kindt R, Legendre P, McGlinn D, et al. vegan: Community Ecology Package [Internet]. 2020. Available from: https://CRAN.R-project.org/package=vegan

32. R Core Team. R: A Language and Environment for Statistical Computing [Internet]. Vienna, Austria: R Foundation for Statistical Computing; 2021. Available from: https://www.R-project.org/

33. Lehto E, Kaartinen NE, Sääksjärvi K, Männistö S, Jallinoja P. Vegetarians and different types of meat eaters among the Finnish adult population from 2007 to 2017. Br J Nutr. 2022 Apr 14;127(7):1060–72.

34. Chen T, Guestrin C. XGBoost: A Scalable Tree Boosting System. In: Proceedings of the 22nd ACM SIGKDD International Conference on Knowledge Discovery and Data Mining [Internet]. New York, NY, USA: Association for Computing Machinery; 2016 [cited 2026 Feb 15]. p. 785–94. (KDD ‘16). Available from: https://dl.acm.org/doi/10.1145/2939672.2939785

35. Kang JW, Khatib LA, Heston MB, Dilmore AH, Labus JS, Deming Y, et al. Gut microbiome compositional and functional features associate with Alzheimer’s disease pathology. Alzheimers Dement J Alzheimers Assoc. 2025 Jul;21(7):e70417.

36. Shvetcov A, Johnson ECB, Winchester LM, Walker KA, Wilkins HM, Thompson TG, et al. APOE ε4 carriers share immune-related proteomic changes across neurodegenerative diseases. Nat Med. 2025 Aug;31(8):2590–601.

37. Allin KH, Tremaroli V, Caesar R, Jensen BAH, Damgaard MTF, Bahl MI, et al. Aberrant intestinal microbiota in individuals with prediabetes. Diabetologia. 2018 Apr;61(4):810–20.

38. Yu Y, Lu J, Sun L, Lyu X, Chang XY, Mi X, et al. Akkermansia muciniphila: A potential novel mechanism of nuciferine to improve hyperlipidemia. Biomed Pharmacother Biomedecine Pharmacother. 2021 Jan;133:111014.

39. Kim S, Lee Y, Kim Y, Seo Y, Lee H, Ha J, et al. Akkermansia muciniphila Prevents Fatty Liver Disease, Decreases Serum Triglycerides, and Maintains Gut Homeostasis. Appl Environ Microbiol. 2020 Mar 18;86(7):e03004–19.

40. Depommier C, Everard A, Druart C, Plovier H, Van Hul M, Vieira-Silva S, et al. Supplementation with Akkermansia muciniphila in overweight and obese human volunteers: a proof-of-concept exploratory study. Nat Med. 2019 Jul;25(7):1096–103.

41. Rao Y, Kuang Z, Li C, Guo S, Xu Y, Zhao D, et al. Gut Akkermansia muciniphila ameliorates metabolic dysfunction-associated fatty liver disease by regulating the metabolism of L-aspartate via gut-liver axis. Gut Microbes. 2021 Dec;13(1):1–19.

42. Zhang X, Shen D, Fang Z, Jie Z, Qiu X, Zhang C, et al. Human gut microbiota changes reveal the progression of glucose intolerance. PloS One. 2013;8(8):e71108.

43. Zapała B, Stefura T, Wójcik-Pędziwiatr M, Kabut R, Bałajewicz-Nowak M, Milewicz T, et al. Differences in the Composition of Gut Microbiota between Patients with Parkinson’s Disease and Healthy Controls: A Cohort Study. J Clin Med. 2021 Dec 3;10(23):5698.

44. Lei W, Cheng Y, Gao J, Liu X, Shao L, Kong Q, et al. Akkermansia muciniphila in neuropsychiatric disorders: friend or foe? Front Cell Infect Microbiol. 2023 Jul 10;13:1224155.

45. Dilmore AH, Martino C, Neth BJ, West KA, Zemlin J, Rahman G, et al. Effects of a ketogenic and low-fat diet on the human metabolome, microbiome, and foodome in adults at risk for Alzheimer’s disease. Alzheimers Dement J Alzheimers Assoc. 2023 Nov;19(11):4805–16.

46. Ou Z, Deng L, Lu Z, Wu F, Liu W, Huang D, et al. Protective effects of Akkermansia muciniphila on cognitive deficits and amyloid pathology in a mouse model of Alzheimer’s disease. Nutr Diabetes. 2020 Apr 22;10(1):12.

47. Ling Z, Zhu M, Yan X, Cheng Y, Shao L, Liu X, et al. Structural and Functional Dysbiosis of Fecal Microbiota in Chinese Patients With Alzheimer’s Disease. Front Cell Dev Biol. 2021 Feb 4;8:634069.

48. Khedr EM, Omeran N, Karam-Allah Ramadan H, Ahmed GK, Abdelwarith AM. Alteration of Gut Microbiota in Alzheimer’s Disease and Their Relation to the Cognitive Impairment. J Alzheimers Dis JAD. 2022;88(3):1103–14.

49. Andreu-Sánchez S, Blanco-Míguez A, Wang D, Golzato D, Manghi P, Heidrich V, et al. Global genetic diversity of human gut microbiome species is related to geographic location and host health. Cell. 2025 Jul 24;188(15):3942-3959.e9.

50. Pei Y, Lu Y, Li H, Jiang C, Wang L. Gut microbiota and intestinal barrier function in subjects with cognitive impairments: a cross-sectional study. Front Aging Neurosci. 2023;15:1174599.

51. Grahnemo L, Nethander M, Coward E, Gabrielsen ME, Sree S, Billod JM, et al. Identification of three bacterial species associated with increased appendicular lean mass: the HUNT study. Nat Commun. 2023 Apr 20;14(1):2250.

52. Naderpoor N, Mousa A, Gomez-Arango LF, Barrett HL, Dekker Nitert M, de Courten B. Faecal Microbiota Are Related to Insulin Sensitivity and Secretion in Overweight or Obese Adults. J Clin Med. 2019 Apr 4;8(4):452.

53. Yang C, Xu J, Xu X, Xu W, Tong B, Wang S, et al. Characteristics of gut microbiota in patients with metabolic associated fatty liver disease. Sci Rep. 2023 Jun 20;13:9988.

54. Y W, Z Z, Jc B, Rl W, Sg O, R W, et al. Antibiotic Use and Subsequent Cognitive Decline and Dementia Risk in Healthy Older Adults. Neurology [Internet]. 2025 Jan 14 [cited 2025 Jul 27];104(1). Available from: https://pubmed.ncbi.nlm.nih.gov/39693592/

55. Martino C, Dilmore AH, Burcham ZM, Metcalf JL, Jeste D, Knight R. Microbiota succession throughout life from the cradle to the grave. Nat Rev Microbiol. 2022 Dec;20(12):707–20.

56. Ghosh TS, Shanahan F, O’Toole PW. The gut microbiome as a modulator of healthy ageing. Nat Rev Gastroenterol Hepatol. 2022 Sep;19(9):565–84.

57. Hirvonen JJ, Lepistö I, Mero S, Kaukoranta SS. First isolation of Dietzia cinnamea from a dog bite wound in an adult patient. J Clin Microbiol. 2012 Dec;50(12):4163–5.

58. Perkin S, Wilson A, Walker D, McWilliams E. Dietzia species pacemaker pocket infection: an unusual organism in human infections. BMJ Case Rep. 2012 Sep 14;2012:bcr1020115011.

59. Muñoz J, Mirelis B, Aragón LM, Gutiérrez N, Sánchez F, Español M, et al. Clinical and microbiological features of nocardiosis 1997-2003. J Med Microbiol. 2007 Apr;56(Pt 4):545–50.

60. La Reau AJ, Strom NB, Filvaroff E, Mavrommatis K, Ward TL, Knights D. Shallow shotgun sequencing reduces technical variation in microbiome analysis. Sci Rep. 2023 May 11;13(1):7668.

61. Chitcharoen S, Sawaswong V, Klomkliew P, Chanchaem P, Payungporn S. Comparative analysis of human gut bacterial microbiota between shallow shotgun metagenomic sequencing and full-length 16S rDNA amplicon sequencing. Biosci Trends. 2025 May 9;19(2):232–42.

62. Govender P, Ghai M. Population-specific differences in the human microbiome: Factors defining the diversity. Gene. 2025 Jan 15;933:148923.

63. Palacios N, Gordon S, Wang T, Burk R, Qi Q, Huttenhower C, et al. Gut microbiome and cognitive function in the Hispanic Community Health Study/Study of Latinos. J Alzheimers Dis JAD. 2025 Nov;108(1):84–97.

64. Karwowska Z, Aasmets O, Metspalu M, Metspalu A, Milani L, Esko T, et al. Effects of data transformation and model selection on feature importance in microbiome classification data. Microbiome. 2025 Jan 4;13(1):2.

65. Forbes JD, Bernstein CN, Tremlett H, Van Domselaar G, Knox NC. A Fungal World: Could the Gut Mycobiome Be Involved in Neurological Disease? Front Microbiol. 2018;9:3249.

