## Supplementary material for "Gut Microbiome and Risk of Dementia –a Prospective, Population-Based Study": Tables_2026-02-15

### Microbiome vs Dementia Tables
2026-01-30

Table 1. Baseline characteristics of the study cohort (n=4,055) according to incident Alzheimer's disease (AD) and all-cause dementia (including AD). P-values represent differences between subjects with and without incident AD or dementia. Welch two sample t-test for continuous and Chi-Square test for categorical variables were applied.

| | Incident AD or dementia | | | | |
|---|---|---|---|---|---|
| | No AD or Dementia (n=3,725) | AD (n=280) | p-value | Dementia (n=330) | p-value |
| Sex (male), % (n) | 46.8 (1,742) | 45.7 (128) | 0.781 | 46.7 (154) | 0.999 |
| Age (years), mean (SD) | 54.4 (8.4) | 64.4 (6.3) | <0.001 | 64.5 (6.4) | <0.001 |
| Number of APOE ε4 alleles | | | | | |
| 0, % (n) | 68.3 (2,543) | 42.5 (119) | | 45.5 (150) | |
| 1, % (n) | 28.9 (1,078) | 43.9 (123) | | 42.7 (141) | |
| 2, % (n) | 2.8 (104) | 13.6 (38) | <0.001 | 11.8 (39) | <0.001 |
| *Education level | | | | | |
| 1, % (n) | 33.9 (1263) | 34.3 (96) | | 35.2 (116) | |
| 2, % (n) | 33.7 (1255) | 28.6 (80) | | 29.1 (96) | |
| 3, % (n) | 32.4 (1207) | 37.1 (104) | 0.146 | 35.7 (118) | 0.211 |
| BMI, kg/m2 (SD) | 27.4 (4.5) | 28.2 (4.2) | 0.003 | 28.3 (4.4) | <0.001 |
| Systolic blood pressure (mmHg), mean (SD) | 139.1 (20.6) | 145.4 (20.4) | <0.001 | 146.9 (20.4) | <0.001 |
| Alcohol consumption, g/week (SD) | 80.5 (127.8) | 60.9 (136.6) | 0.021 | 59.5 (136.7) | 0.008 |

| | | | | | |
|---|---|---|---|---|---|
| Smoking | | | | | |
| Never, % (n) | 51.2 (1906) | 61.1 (171) | | 60.3 (199) | |
| Ex-smoker Cessation, % (n) | 26.0 (967) | 22.9 (64) | | 23.6 (78) | |
| Current, % (n) | 22.9 (852) | 16.1 (45) | 0.004 | 16.1 (53) | 0.003 |
| Anti-Hypertensive Medication, % (n) | 19.1 (711) | 32.5 (91) | <0.001 | 33.9 (112) | <0.001 |
| Gut microbiome altering drugs**, % (n) | 47.6 (1774) | 63.6 (178) | 0.001 | 65.6 (217) | <0.001 |
| Prevalent | | | | | |
| Diabetes, n (%) | 4.3 (159) | 7.1 (20) | 0.036 | 7.0 (23) | 0.033 |
| CVD, % (n) | 3.7 (139) | 6.8 (19) | <0.001 | 7.3 (24) | 0.003 |

Abbrevations: BMI – Body Mass Index, CVD – Cardiovascular Disease.

*Education level, self-reported duration of education further graded into tertiles and adjusted for the birth year.

**Gut microbiome altering drugs, see supplementary Table 1.

Table 2. Associations between all taxa detected in more than 1% of subjects and Alzheimer’s disease or Dementia. P-values were FDR corrected (q-value) and only taxa with a q-values<0.05 for AD, dementia or both are presented.

| | Alzheimer’s Disease | | | Dementia | | |
|---|---|---|---|---|---|---|
| Predictor | HR (95%CI) | P-value | q-value | HR (95%CI) | P-value | q-value |
| **Genera** | | | | | | |
| *Dietzia* | 1.12 (0.99-1.28) | 0.082 | 0.302 | 1.17 (1.09-1.25) | <0.001 | 0.013 |
| **Species** | | | | | | |
| *Brevibacterium epidermidis* | 1.09 (1.01-1.18) | 0.021 | 0.220 | 1.08 (1.05-1.11) | <0.001 | 0.001 |
| *Liquorilactobacillus vini* | 1.17 (1.08-1.26) | <0.001 | 0.220 | 1.17 (1.09-1.26) | <0.001 | 0.019 |

Results are based on the Fine-Gray model adjusted for Sex, Number of APOE ε4 alleles, Education level, BMI, Systolic blood pressure, Alcohol consumption, Smoking status, Usage of Anti-Hypertensive Medication, Gut microbiome altering drugs, Prevalent Diabetes, Prevalent CVD. Age was used as the time scale.

Table 3. Associations between phyla and orders linked to APOE genotype and Alzheimer's disease or Dementia. Results are ordered by FDR corrected p-values (q-values). Family, Genera and Species level results are presented in the Supplement.

| | Alzheimer's Disease | | | Dementia | | |
|---|---|---|---|---|---|---|
| **Phylum** | HR (95%CI) | P-value | q-value | HR (95%CI) | P-value | q-value |
| *Verrucomicrobiota* | 1.19 (1.02-1.39) | 0.027 | 0.054 | 1.20 (1.03-1.39) | 0.020 | 0.040 |
| *Campylobacterota* | 0.94 (0.81-1.09) | 0.399 | 0.399 | 0.92 (0.80-1.05) | 0.210 | 0.210 |
| **Orders** | | | | | | |
| *Verrucomicrobiales* | 1.22 (1.05-1.41) | 0.010 | 0.060 | 1.21 (1.05-1.40) | 0.009 | 0.054 |
| *Tissierellales* | 1.20 (1.01-1.42) | 0.033 | 0.099 | 1.15 (0.98-1.34) | 0.085 | 0.255 |
| *Campylobacterales* | 0.94 (0.82-1.09) | 0.431 | 0.681 | 0.92 (0.80-1.05) | 0.226 | 0.452 |
| *Xenobiales* | 0.94 (0.81-1.10) | 0.454 | 0.681 | 0.93 (0.78-1.10) | 0.382 | 0.573 |
| *Veillonellales* | 1.03 (0.89-1.19) | 0.660 | 0.792 | 1.04 (0.91-1.19) | 0.575 | 0.690 |
| *RFP12* | 1.00 (0.86-1.16) | 0.976 | 0.976 | 1.03 (0.89-1.18) | 0.730 | 0.730 |

Results are based on the Fine-Gray model adjusted for Sex, APOE genotype (0 vs 1-2 ε alleles), Education level, BMI, Systolic blood pressure, Alcohol consumption, Smoking status, Usage of Anti-Hypertensive Medication, Gut microbiome altering drugs, Prevalent Diabetes, Prevalent CVD. Age was used as the time scale.
