## Supplements_2026-02-15 for "Gut Microbiome and Risk of Dementia –a Prospective, Population-Based Study"

Supplementary Table 1. List of potentially gut microbiome altering drugs according to ATC classification.

| | ATC classification |
|---|---|
| A | Alimentary tract and metabolism |
| B | Blood and blood forming organs |
| C | Cardiovascular system |
| G | Genito-urinary system and sex hormones |
| H | Systemic hormonal preparations, excluding sex hormones and insulins |
| J | Anti-infective for systemic use |
| L | Antineoplastic and immunomodulating agents |
| N | Nervous system |
| P | Antiparasitic products, insecticides and repellents |

www.who.int/tools/atc-ddd-toolkit/atc-classification

Supplementary Table 2. Hazard ratios (HR) for incident AD and dementia based on Fine-Gray subdistribution model.

| | | Alzheimer's Disease | | Dementia | |
|---|---|---|---|---|---|
| | | HR (95%CI) | P-value | HR (95%CI) | P-value |
| Sex (male as reference) | | 0.74 (0.53-1.03) | 0.075 | 0.70 (0.51-0.95) | 0.024 |
| Number of ApoE ε4 alleles | | 2.77 (2.25-3.41) | <0.001 | 2.62 (2.15-3.18) | <0.001 |
| Education level* | 1 | ref | | ref | |
| | 2 | 0.75 (0.52-1.06) | 0.102 | 0.76 (0.55-1.05) | 0.101 |
| | 3 | 0.96 (0.69-1.35) | 0.817 | 0.95 (0.69-1.30) | 0.749 |
| BMI (kg/m2) | | 1.00 (0.97-1.03) | 0.964 | 1.00 (0.97-1.02) | 0.859 |
| Systolic blood pressure (mmHg) | | 1.00 (0.99-1.00) | 0.498 | 1.00 (0.99-1.01) | 0.998 |
| Alcohol Consumption (doses/x) | | 1.00 (1.00-1.00) | 0.908 | 1.00 (1.00-1.00) | 0.868 |
| Smoking | Never | ref | | ref | |
| | Cessation | 0.94 (0.65-1.35) | 0.738 | 0.99 (0.71-1.39) | 0.955 |
| | Current | 1.06 (0.71-1.56) | 0.786 | 1.05 (0.73-1.51) | 0.799 |
| Anti-Hypertensive Medication | | 1.23 (0.86-1.76) | 0.264 | 1.26 (0.90-1.75) | 0.174 |
| Gut microbiome altering drugs** | | 1.20 (0.84-1.71) | 0.319 | 1.22 (0.87-1.70) | 0.245 |
| Prevalent | Diabetes | 1.04 (0.66-1.64) | 0.859 | 1.06 (0.70-1.60) | 0.796 |
| | CVD | 1.02 (0.68-1.52) | 0.939 | 1.11 (0.77-1.60) | 0.561 |

Abbrevations: BMI – Body Mass Index, CVD – Cardio Vascular Disease.

*Education level, self-reported duration of education further graded into tertiles and adjusted for the birth year.

**Gut microbiome altering drugs purchase of within 0-4 months prior to the study baseline. See supplementary Table 1.

Supplementary Table 3. Association (p-term) of dbRDA with Alzheimer's and dementia risk.

| | Phylum | Genus | Species |
|---|---|---|---|
| Dementia | 0.049 | 0.009 | 0.007 |
| Alzheimer's disease | 0.146 | 0.089 | 0.058 |

dbRDA results are based on logistic regression model adjusted for Age, Sex, Number of APOE ε4 alleles, Education level, BMI, Systolic blood pressure, Alcohol consumption, Smoking status, Usage of Anti-Hypertensive Medication, Gut microbiome altering drugs, Prevalent Diabetes, Prevalent CVD.

Supplementary Table 4. Association between the first three principal components (PC) of β-diversity, based on Aitchison distance, and Alzheimer’s disease or dementia.

| | Alzheimer’s Disease HR (95%CI) | P-value | Dementia HR (95%CI) | P-value |
|---|---|---|---|---|
| PC 1 | 1.08 (0.97-1.20) | 0.161 | 1.01 (0.93-1.11) | 0.784 |
| PC 2 | 0.93 (0.83-1.04) | 0.201 | 0.93 (0.84-1.04) | 0.210 |
| PC 3 | 0.97 (0.87-1.08) | 0.608 | 0.96 (0.87-1.06) | 0.415 |

Results are based on the Fine-Gray model adjusted for Sex, Number of APOE ε4 alleles, Education level, BMI, Systolic blood pressure, Alcohol consumption, Smoking status, Usage of Anti-Hypertensive Medication, Gut microbiome altering drugs, Prevalent Diabetes, Prevalent CVD. Age was used as the time scale.

Supplementary Table 5. Association between the first three microbial principal components (PC) and Alzheimer’s disease or dementia.

| | Alzheimer’s Disease HR (95%CI) | P-value | Dementia HR (95%CI) | P-value |
|---|---|---|---|---|
| PC 1 | 0.90 (0.80-1.01) | 0.066 | 0.93 (0.84-1.03) | 0.171 |
| PC 2 | 1.01 (0.91-1.12) | 0.849 | 1.05 (0.96-1.15) | 0.308 |
| PC 3 | 1.03 (0.92-1.14) | 0.645 | 0.98 (0.89-1.08) | 0.685 |

Results are based on the Fine-Gray model adjusted for Sex, Number of APOE ε4 alleles, Education level, BMI, Systolic blood pressure, Alcohol consumption, Smoking status, Usage of Anti-Hypertensive Medication, Gut microbiome altering drugs, Prevalent Diabetes, Prevalent CVD. Age was used as the time scale.

Supplementary Table 6. Associations between candidate microbes and Incident Alzheimer's Disease and dementia.

| | | Previously detected abundance in Alzheimer's Disease (↓ = lower and ↑= higher abundance) and study reference number | Alzheimer's Disease HR (95%CI) | P-value | Dementia HR (95%CI) | P-value |
|---|---|---|---|---|---|---|
| Phylum | *Actinobacteria* | ↓ (1,2) | 0.97 (0.88-1.07) | 0.562 | 0.97 (0.88-1.07) | 0.555 |
| | *Bacteroidetes* | ↑ (1,2), ↓ (3,4) | 0.95 (0.86-1.04) | 0.277 | 0.94 (0.86-1.02) | 0.152 |
| | *Firmicutes** | ↓ (1,2,3), ↑ (4) | 0.93 (0.83-1.05) | 0.234 | 0.94 (0.84-1.05) | 0.243 |
| | *Proteobacteria* | ↑ (1,3) | 0.98 (0.88-1.08) | 0.644 | 1.02 (0.93-1.13) | 0.661 |
| Family | *Bacteroidaceae* | ↑ (1) | 0.96 (0.87-1.06) | 0.462 | 0.94 (0.85-1.03) | 0.173 |
| | *Bifidobacteriaceae* | ↓ (1) | 1.00 (0.90-1.11) | 0.992 | 0.99 (0.90-1.09) | 0.838 |
| | *Clostridiaceae** | ↓ (1) | 0.95 (0.84-1.07) | 0.365 | 0.93 (0.83-1.04) | 0.213 |
| | *Erysipelotrichaceae* | ↓ (1) | 1.00 (0.93-1.08) | 0.992 | 1.00 (0.93-1.07) | 0.894 |
| | *Lactobacillaceae* | ↑(5) | 1.07 (0.96-1.21) | 0.232 | 1.04 (0.94-1.16) | 0.432 |
| | *Peptostreptococcaceae** | ↓ (1) | 1.08 (0.97-1.21) | 0.156 | 1.02 (0.92-1.13) | 0.688 |
| | *Rikenellaceae* | ↑ (1) | 1.08 (0.96-1.20) | 0.207 | 1.05 (0.95-1.17) | 0.337 |
| | *Ruminococcaceae* | ↓ (1) | 1.03 (0.92-1.16) | 0.562 | 0.97 (0.88-1.08) | 0.614 |
| Genus | *Adlercreutzia** | ↓ (1) | 0.97 (0.87-1.09) | 0.626 | 0.97 (0.87-1.07) | 0.492 |

| | | | | | |
|---|---|---|---|---|---|
| *Alistipes** | ↑ (1), ↓ (4) | 1.06 (0.94-1.19) | 0.351 | 1.05 (0.94-1.17) | 0.407 |
| *Bacteroides** | ↑ (1) | 0.97 (0.87-1.09) | 0.635 | 0.96 (0.86-1.06) | 0.406 |
| *Bifidobacterium** | ↓(2), ↑ (4) | 0.98 (0.87-1.09) | 0.688 | 0.97 (0.88-1.08) | 0.626 |
| *Bilophila* | ↑ (1) | 1.11 (0.99-1.24) | 0.081 | 1.10 (0.99-1.23) | 0.070 |
| *Blautia** | ↑ (1,4) | 0.93 (0.83-1.06) | 0.279 | 0.94 (0.85-1.05) | 0.313 |
| *Collinsella* | ↑(6) | 1.00 (0.90-1.12) | 0.971 | 1.01 (0.91-1.12) | 0.817 |
| *Clostridium** | ↓ (1) | 0.96 (0.85-1.08) | 0.524 | 0.93 (0.83-1.05) | 0.232 |
| *Dialister* | ↓ (1) | 1.08 (0.97-1.21) | 0.147 | 1.09 (0.99-1.21) | 0.086 |
| *Dorea** | ↑ (4) | 0.92 (0.83-1.03) | 0.167 | 0.90 (0.81-0.99) | 0.038 |
| *Escherichia** | ↑ (4) | 1.01 (0.90-1.13) | 0.923 | 1.05 (0.94-1.16) | 0.406 |
| *Eubacterium** | ↓ (6) | 0.98 (0.87-1.09) | 0.672 | 0.96 (0.87-1.06) | 0.440 |
| *Faecalibacterium* | ↓(5) | 1.07 (0.967-1.20) | 0.183 | 1.04 (0.95-1.14) | 0.417 |
| *Lactobacillus* | ↑ (4) | 1.10 (0.98-1.24) | 0.120 | 1.06 (0.95-1.18) | 0.334 |
| *Parabacteroides** | ↓ (4) | 1.09 (0.97-1.24) | 0.158 | 1.09 (0.97-1.22) | 0.160 |
| *Paraprevotella* | ↓ (4) | 0.99 (0.90-1.09) | 0.822 | 0.99 (0.90-1.09) | 0.880 |
| *Phascolarctobacterium* * | ↑ (1) | 0.99 (0.89-1.10) | 0.817 | 0.97 (0.87-1.07) | 0.525 |

| | | | | | |
|---|---|---|---|---|---|
| *Ruminiclostridium* | ↑↓(5) | 0.91 (0.81-1.02) | 0.118 | 0.94 (0.84-1.04) | 0.213 |
| *Streptococcus* | ↑ (4) | 0.99 (0.89-1.10) | 0.838 | 0.98 (0.89-1.09) | 0.747 |
| *Sutterella* | ↓ (4) | 1.02 (0.92-1.14) | 0.688 | 1.01 (0.92-1.12) | 0.804 |
| *Turicibacter* | ↓ (1) | 1.06 (0.95-1.18) | 0.274 | 1.02 (0.92-1.12) | 0.719 |
| *Veillonella** | ↑(6) | 1.10 (0.99-1.22) | 0.075 | 1.07 (0.97-1.18) | 0.174 |

Results are based on the Fine-Gray model adjusted for Sex, Number of APOE ε4 alleles, Education level, BMI, Systolic blood pressure, Alcohol consumption, Smoking status, Usage of Anti-Hypertensive Medication, Gut microbiome altering drugs, Prevalent Diabetes, Prevalent CVD. Age was used as the time scale.

*Based on SHOGUN v. 1.0.5 pipeline, other results are based on Greengenes2 reference tree.

References:


1. Vogt NM, Kerby RL, Dill-McFarland KA, Harding SJ, Merluzzi AP, Johnson SC, ym. Gut microbiome alterations in Alzheimer's disease. Sci Rep. 19. Dec 2017;7(1):13537.
2. Khedr EM, Omeran N, Karam-Allah Ramadan H, Ahmed GK, Abdelwarith AM. Alteration of Gut Microbiota in Alzheimer's Disease and Their Relation to the Cognitive Impairment. J Alzheimers Dis JAD. 2022;88(3):1103–14.
3. Liu P, Wu L, Peng G, Han Y, Tang R, Ge J, ym. Altered microbiomes distinguish Alzheimer's disease from amnestic mild cognitive impairment and health in a Chinese cohort. Brain Behav Immun. Aug 2019;80:633–43.
4. Li B, He Y, Ma J, Huang P, Du J, Cao L, ym. Mild cognitive impairment has similar alterations as Alzheimer's disease in gut microbiota. Alzheimers Dement J Alzheimers Assoc. Oct 2019;15(10):1357–66.
5. Ning J, Huang SY, Chen SD, Zhang YR, Huang YY, Yu JT. Investigating Casual Associations Among Gut Microbiota, Metabolites, and Neurodegenerative Diseases: A Mendelian Randomization Study. J Alzheimers Dis JAD. 2022;87(1):211–22.
6. Cammann D, Lu Y, Cummings MJ, Zhang ML, Cue JM, Do J, ym. Genetic correlations between Alzheimer's disease and gut microbiome genera. Sci Rep. 31. March 2023;13:5258.

Supplementary Table 7. Associations between *species* and Dementia. Only FDR–corrected p-values (q-value < 0.05) are presented. Analysis is adjusted for clinical risk factors and APOE carrier status (0 vs 1-2 alleles) with 10 years follow-up time restriction. No associations (q-value < 0.05) were observed between *species* and AD in this analysis.

| Specie | HR (95%CI) | P-value | q-value |
|---|---|---|---|
| *Brevibacterium epidermidis* | 1.15 (1.11-1.19) | <0.001 | <0.001 |
| *Corynebacterium stationis* | 1.11 (1.07-1.15) | <0.001 | <0.001 |
| *Leucobacter sp000980875* | 1.14 (1.09-1.19) | <0.001 | <0.001 |
| *Dietzia cinnamea* | 1.19 (1.11-1.26) | <0.001 | <0.001 |
| *Corynebacterium efficiens* | 1.11 (1.06-1.16) | <0.001 | 0.006 |
| *Mobilicoccus pelagius* | 1.17 (1.08-1.26) | <0.001 | 0.041 |
| *Acinetobacter bouvetii* | 1.15 (1.07-1.23) | <0.001 | 0.041 |

Results are based on the Fine-Gray model adjusted for Sex, APOE carrier (0 vs 1-2 alleles), Education level, BMI, Systolic blood pressure, Alcohol consumption, Smoking status, Usage of Anti-Hypertensive Medication, Gut microbiome altering drugs, Prevalent Diabetes, Prevalent CVD. Age was used as the time scale.

Supplementary Table 8. Statistically significant (p<0.05) linear associations between APOE genotype and microbe abundances.

| **Phylum** | P-value | Beta | SE |
|---|---|---|---|
| *Campylobacterota* | 0.022 | -0.065 | 0.028 |
| *Verrucomicrobiota* | 0.003 | 0.082 | 0.028 |
| **Order** | | | |
| *Campylobacterales* | 0.023 | -0.064 | 0.028 |
| *RFP12* | 0.045 | 0.056 | 0.028 |
| *Tissierellales* | 0.032 | -0.060 | 0.028 |
| *Veillonellales* | 0.040 | 0.058 | 0.028 |
| *Verrucomicrobiales* | 0.001 | 0.092 | 0.028 |
| *Xenobiales* | 0.032 | 0.056 | 0.026 |
| **Family** | | | |
| *Akkermansiaceae* | 0.001 | 0.091 | 0.028 |
| *Aneurinibacillaceae* | 0.037 | -0.060 | 0.029 |
| *Beijerinckiaceae* | 0.030 | -0.061 | 0.028 |
| *CAG.552* | 0.048 | 0.056 | 0.028 |
| *CAG.826* | 0.006 | 0.078 | 0.028 |
| *Campylobacteraceae* | 0.029 | -0.062 | 0.028 |
| *Desulfofarciminaceae* | 0.022 | 0.065 | 0.028 |
| *Dialisteraceae* | 0.028 | 0.063 | 0.028 |
| *Marinococcaceae* | 0.017 | 0.068 | 0.028 |
| *PHET01* | 0.047 | 0.055 | 0.028 |
| *Pseudomonadaceae* | 0.038 | -0.058 | 0.028 |
| *Salinivirgaceae* | 0.042 | 0.058 | 0.028 |
| *Salisediminibacteriaceae* | 0.024 | 0.063 | 0.028 |
| *Sulfurimonadaceae* | 0.025 | 0.064 | 0.029 |
| *Tissierellaceae* | 0.033 | -0.059 | 0.028 |
| *UBA10428* | 0.048 | 0.057 | 0.029 |
| *UBA1067* | 0.046 | 0.056 | 0.028 |
| *WCHB1.69* | 0.020 | -0.066 | 0.028 |
| *Xenobiaceae* | 0.038 | 0.054 | 0.026 |
| **Genus** | | | |
| *Acetatifactor* | 0.010 | -0.072 | 0.028 |
| *Advenella* | 0.018 | 0.071 | 0.030 |
| *Akkermansia* | 0.001 | 0.092 | 0.028 |
| *Bisgaardia* | 0.009 | 0.076 | 0.029 |
| *Bosea* | 0.038 | -0.059 | 0.028 |
| *Brochothrix* | 0.009 | 0.074 | 0.029 |
| *Bruticola* | 0.038 | 0.055 | 0.026 |
| *Cloacibacterium* | 0.029 | 0.063 | 0.029 |
| *Desulfofarcimen* | 0.021 | 0.065 | 0.028 |
| *Dialister* | 0.028 | 0.063 | 0.028 |
| *Dolosicoccus* | 0.014 | 0.068 | 0.028 |
| *DSSC01* | 0.047 | 0.055 | 0.028 |
| *DUUS01* | 0.035 | -0.059 | 0.028 |
| *F0540* | 0.032 | 0.062 | 0.029 |
| *Fundidesulfovibrio* | 0.025 | -0.064 | 0.028 |
| *JAAAOM01* | 0.046 | 0.057 | 0.029 |
| *JAAYUG01* | 0.050 | 0.051 | 0.026 |
| *Lichenihabitans* | 0.049 | 0.054 | 0.028 |
| *Marinococcus* | 0.019 | 0.067 | 0.029 |
| *Methanofastidiosum* | 0.006 | 0.078 | 0.028 |

| | | | |
|---|---|---|---|
| *Methyloterricola* | 0.035 | 0.059 | 0.028 |
| *Neobacillus* | 0.038 | -0.059 | 0.028 |
| *Olegusella* | 0.017 | -0.068 | 0.029 |
| *Oliverpabstia* | 0.039 | -0.059 | 0.028 |
| *Parasutterella* | 0.046 | -0.057 | 0.028 |
| *Proteus* | 0.046 | 0.057 | 0.028 |
| *Psychrosphaera* | 0.033 | 0.060 | 0.028 |
| *Roseburia* | 0.038 | -0.057 | 0.027 |
| *RUG13038* | 0.043 | -0.057 | 0.028 |
| *Salimicrobium* | 0.014 | 0.069 | 0.028 |
| *Salinivirga* | 0.041 | 0.058 | 0.028 |
| *Schleiferilactobacillus* | 0.015 | -0.070 | 0.028 |
| *Sulfuricurvum* | 0.037 | 0.059 | 0.028 |
| *UBA3789* | 0.026 | -0.063 | 0.028 |
| *UBA4951* | 0.004 | 0.082 | 0.028 |
| *UPXZ01* | 0.048 | -0.056 | 0.028 |
| *VFKE01* | 0.031 | 0.062 | 0.029 |
| *Winkia* | 0.016 | -0.067 | 0.028 |
| **Species** | | | |
| *Acetatifactor sp900066565* | 0.009 | -0.073 | 0.028 |
| *Actinomyces slackii* | 0.034 | 0.058 | 0.028 |
| *Advenella mimigardefordensis* | 0.012 | 0.075 | 0.030 |
| *Aeromicrobium choanae* | 0.036 | 0.063 | 0.030 |
| *Aggregatibacter actinomycetemcomitans* | 0.038 | 0.059 | 0.028 |
| *Akkermansia glycaniphila* | 0.037 | 0.059 | 0.028 |
| *Alloscardovia macacae* | 0.034 | 0.059 | 0.028 |
| *Arcticibacter svalbardensis* | 0.045 | 0.054 | 0.027 |
| *Bifidobacterium crudilactis* | 0.030 | 0.061 | 0.028 |
| *Bifidobacterium psychraerophilum* | 0.043 | 0.057 | 0.028 |
| *Bifidobacterium swidsinskii* | 0.049 | 0.056 | 0.028 |
| *Bisgaardia hudsonensis* | 0.009 | 0.076 | 0.029 |
| *Brevibacterium senegalense* | 0.003 | 0.079 | 0.027 |
| *Brochothrix campestris* | 0.027 | 0.064 | 0.029 |
| *Brochothrix thermosphacta* | 0.030 | 0.062 | 0.029 |
| *Bruticola papionis* | 0.033 | 0.056 | 0.026 |
| *CAG 313 sp000433035* | 0.047 | 0.056 | 0.028 |
| *Cellulosilyticum ruminicola* | 0.022 | 0.065 | 0.029 |
| *Cloacibacterium normanense* | 0.029 | 0.063 | 0.029 |
| *Corynebacterium simulans* | 0.024 | 0.065 | 0.029 |
| *Dermacoccus profundi* | 0.035 | 0.061 | 0.029 |
| *Desulfofarcimen acetoxidans* | 0.021 | 0.065 | 0.028 |
| *Dialister invisus* | 0.036 | 0.060 | 0.028 |
| *Dolosicoccus paucivorans* | 0.014 | 0.068 | 0.028 |
| *DSSC01 sp011331305* | 0.041 | 0.056 | 0.028 |
| *DUUS01 sp012841465* | 0.036 | -0.058 | 0.028 |
| *Dwaynesavagella sp000283555* | 0.008 | 0.077 | 0.029 |
| *Dysgonomonas capnocytophagoides* | 0.037 | 0.062 | 0.030 |
| *Ectobacillus funiculus* | 0.047 | 0.058 | 0.029 |
| *Enteromonas sp002472275* | 0.041 | 0.058 | 0.028 |
| *F0540 sp000466585* | 0.029 | 0.064 | 0.029 |
| *FEB 22 sp003105195* | 0.040 | 0.059 | 0.029 |
| *Fimenecus sp900315985* | 0.029 | -0.062 | 0.028 |
| *Flavobacterium cauense* | 0.035 | 0.058 | 0.028 |
| *Fundidesulfovibrio putealis* | 0.024 | -0.064 | 0.028 |

| | | | |
|---|---|---|---|
| *Ilyobacter polytropus* | 0.036 | 0.060 | 0.028 |
| *JAAAOM01 sp009908855* | 0.044 | 0.058 | 0.029 |
| *JAAYUG01 sp012517825* | 0.045 | 0.052 | 0.026 |
| *Lacticaseibacillus pantheris* | 0.046 | 0.059 | 0.030 |
| *Lactiplantibacillus xiangfangensis* | 0.028 | -0.062 | 0.028 |
| *Lactobacillus melliventris* | 0.024 | 0.058 | 0.026 |
| *Lentilactobacillus farraginis* | 0.034 | -0.059 | 0.028 |
| *Lentilactobacillus hilgardii* | 0.003 | -0.085 | 0.028 |
| *Ligilactobacillus agilis* | 0.004 | 0.080 | 0.028 |
| *Ligilactobacillus animalis* | 0.008 | 0.074 | 0.028 |
| *Ligilactobacillus apodemi* | 0.038 | 0.057 | 0.028 |
| *Ligilactobacillus equi* | 0.033 | 0.061 | 0.028 |
| *Ligilactobacillus murinus* | 0.008 | 0.073 | 0.027 |
| *Liquorilactobacillus uvarum* | 0.011 | -0.072 | 0.028 |
| *Marinococcus luteus* | 0.017 | 0.068 | 0.029 |
| *Methanofastidiosum sp001587595* | 0.006 | 0.078 | 0.028 |
| *Methyloterricola oryzae* | 0.025 | 0.063 | 0.028 |
| *Neobacillus thermocopriae* | 0.013 | 0.063 | 0.025 |
| *Nocardia carnea* | 0.019 | 0.069 | 0.029 |
| *Olegusella massiliensis* | 0.018 | -0.068 | 0.029 |
| *Oliverpabstia intestinalis* | 0.040 | -0.059 | 0.028 |
| *Paracoccus yeei* | 0.048 | 0.052 | 0.026 |
| *Parasutterella excrementihominis* | 0.016 | -0.068 | 0.028 |
| *Pediococcus inopinatus* | 0.012 | -0.071 | 0.028 |
| *Pediococcus stilesii* | 0.035 | 0.060 | 0.028 |
| *Pedobacter insulae* | 0.005 | 0.081 | 0.029 |
| *Pedobacter sp001027745* | 0.017 | 0.070 | 0.030 |
| *Prevotella micans* | 0.018 | -0.067 | 0.028 |
| *Prevotella oris* | 0.032 | -0.061 | 0.028 |
| *Prevotella sp000688375* | 0.029 | -0.062 | 0.028 |
| *Propionibacterium cyclohexanicum* | 0.011 | 0.071 | 0.028 |
| *Proteus mirabilis* | 0.030 | 0.062 | 0.028 |
| *Psychrosphaera saromensis* | 0.030 | 0.061 | 0.028 |
| *Ralstonia pseudosolanacearum* | 0.046 | 0.059 | 0.030 |
| *Roseburia inulinivorans* | 0.002 | -0.086 | 0.028 |
| *Rothia aeria* | 0.047 | -0.056 | 0.028 |
| *RUG13038 sp902789425* | 0.050 | -0.055 | 0.028 |
| *Salimicrobium jeotgali* | 0.010 | 0.073 | 0.028 |
| *Salinivibrio socompensis* | 0.044 | 0.048 | 0.024 |
| *Salinivirga cyanobacteriivorans* | 0.041 | 0.058 | 0.028 |
| *Schleiferilactobacillus perolens* | 0.047 | -0.057 | 0.029 |
| *Staphylococcus capitis* | 0.040 | 0.059 | 0.029 |
| *Sulfuricurvum sp002327465* | 0.014 | 0.069 | 0.028 |
| *UBA3789 sp002369415* | 0.041 | -0.058 | 0.029 |
| *UBA3789 sp902780585* | 0.049 | -0.055 | 0.028 |
| *UBA3947 sp900095445* | 0.016 | 0.070 | 0.029 |
| *UBA4951 sp900542595* | 0.001 | 0.092 | 0.028 |
| *UPXZ01 sp900498215* | 0.034 | -0.060 | 0.028 |
| *VFKE01 sp009885695* | 0.029 | 0.062 | 0.029 |
| *Vibrio sp001506075* | 0.023 | 0.060 | 0.026 |
| *Winkia sp002849225* | 0.033 | -0.059 | 0.028 |

The results are adjusted for Age, Sex, Education level, BMI, Systolic blood pressure, Alcohol consumption, Smoking status, Usage of Anti-Hypertensive Medication, Gut microbiome altering drugs, Prevalent Diabetes and Prevalent CVD.

Supplementary Table 9. Associations between families linked to APOE genotype and Alzheimer’s disease or dementia. The results are ordered based on FDR–corrected p-values (q-value).

| | Alzheimer’s Disease | | | | Dementia | | |
|---|---|---|---|---|---|---|---|
| Predictor for Alzheimer’s Disease | HR (95%CI) | P-value | q-value | Predictor for Dementia | HR (95%CI) | P-value | q-value |
| *Akkermansiaceae* | 1.22 (1.05-1.41) | 0.010 | 0.190 | *Akkermansiaceae* | 1.21 (1.05-1.40) | 0.008 | 0.152 |
| *WCHB1.69* | 1.21 (1.02-1.44) | 0.032 | 0.291 | *WCHB1.69* | 1.13 (0.99-1.28) | 0.060 | 0.507 |
| *Tissierellaceae* | 1.18 (1.00-1.39) | 0.046 | 0.291 | *Marinococcaceae* | 0.82 (0.66-1.02) | 0.080 | 0.507 |
| *Marinococcaceae* | 0.81 (0.65-1.02) | 0.080 | 0.327 | *Tissierellaceae* | 1.12 (0.96-1.30) | 0.146 | 0.614 |
| *Salisediminibacteriaceae* | 0.85 (0.70-1.02) | 0.086 | 0.327 | *Campylobacteraceae* | 0.91 (0.80-1.04) | 0.185 | 0.614 |
| *Aneurinibacillaceae* | 0.86 (0.67-1.10) | 0.223 | 0.706 | *Aneurinibacillaceae* | 0.88 (0.72-1.07) | 0.194 | 0.614 |
| *Campylobacteraceae* | 0.93 (0.81-1.07) | 0.296 | 0.772 | *Salisediminibacteriaceae* | 0.91 (0.77-1.08) | 0.283 | 0.768 |
| *CAG.826* | 1.07 (0.93-1.23) | 0.325 | 0.772 | *Sulfurimonadaceae* | 0.91 (0.74-1.12) | 0.377 | 0.788 |
| *CAG.552* | 1.06 (0.93-1.21) | 0.404 | 0.777 | *Xenobiaceae* | 0.93 (0.79-1.10) | 0.407 | 0.788 |
| *Sulfurimonadaceae* | 0.93 (0.79-1.10) | 0.409 | 0.777 | *CAG.552* | 1.06 (0.93-1.20) | 0.415 | 0.788 |
| *Xenobiaceae* | 0.95 (0.81-1.10) | 0.465 | 0.803 | *PHET01* | 0.93 (0.75-1.13) | 0.457 | 0.789 |
| *Pseudomonadaceae* | 0.95 (0.82-1.11) | 0.527 | 0.829 | *Dialisteraceae* | 1.05 (0.92-1.20) | 0.500 | 0.792 |
| *PHET01* | 0.95 (0.79-1.14) | 0.567 | 0.829 | *CAG.826* | 1.03 (0.91-1.18) | 0.608 | 0.832 |
| *Dialisteraceae* | 1.03 (0.90-1.19) | 0.639 | 0.867 | *Desulfofarciminaceae* | 0.96 (0.80-1.14) | 0.613 | 0.832 |
| *Desulfofarciminaceae* | 0.98 (0.84-1.15) | 0.824 | 0.987 | *Pseudomonadaceae* | 0.97 (0.85-1.11) | 0.669 | 0.847 |
| *UBA10428* | 1.01 (0.89-1.15) | 0.847 | 0.987 | *UBA1067* | 1.03 (0.89-1.18) | 0.719 | 0.854 |
| *Salinivirgaceae* | 1.00 (0.87-1.16) | 0.967 | 0.987 | *Salinivirgaceae* | 0.98 (0.85-1.13) | 0.818 | 0.914 |
| *UBA1067* | 1.00 (0.86-1.16) | 0.982 | 0.987 | *Beijerinckiaceae* | 1.01 (0.87-1.17) | 0.901 | 0.951 |
| *Beijerinckiaceae* | 1.00 (0.86-1.17) | 0.987 | 0.987 | *UBA10428* | 1.00 (0.87-1.14) | 0.951 | 0.951 |

Results are based on the Fine-Gray model adjusted for Sex, APOE genotype (0 vs 1-2 ε alleles), Education level, BMI, Systolic blood pressure, Alcohol consumption, Smoking status, Usage of Anti-Hypertensive Medication, Gut microbiome altering drugs, Prevalent Diabetes, Prevalent CVD. Age was used as the time scale.

Supplementary Table 10. Associations between genera linked to APOE genotype and Alzheimer's disease or dementia. The results are ordered based on FDR–corrected p-values (q-value).

| Alzheimer's Disease | | | | Dementia | | | |
|---|---|---|---|---|---|---|---|
| Predictor | HR (95%CI) | P-value | q-value | Predictor | HR (95%CI) | P-value | q-value |
| *Akkermansia* | 1.21 (1.05-1.41) | 0.010 | 0.228 | *Akkermansia* | 1.21 (1.05-1.40) | 0.008 | 0.266 |
| *JAAYUG01* | 0.77 (0.63-0.94) | 0.012 | 0.228 | *JAAYUG01* | 0.77 (0.63-0.95) | 0.014 | 0.266 |
| *Marinococcus* | 0.81 (0.66-0.99) | 0.036 | 0.456 | *Marinococcus* | 0.84 (0.70-1.01) | 0.069 | 0.836 |
| *Winkia* | 0.74 (0.53-1.03) | 0.075 | 0.713 | *Winkia* | 0.78 (0.59-1.04) | 0.088 | 0.836 |
| *Fundidesulfovibrio* | 0.84 (0.67-1.05) | 0.127 | 0.804 | *Fundidesulfovibrio* | 0.89 (0.76-1.04) | 0.146 | 0.871 |
| *Salimicrobium* | 0.90 (0.79-1.03) | 0.127 | 0.804 | *Salimicrobium* | 0.88 (0.74-1.05) | 0.154 | 0.871 |
| *Advenella* | 0.91 (0.79-1.05) | 0.190 | 0.990 | *Advenella* | 1.08 (0.97-1.21) | 0.170 | 0.871 |
| *Acetatifactor* | 1.08 (0.96-1.22) | 0.214 | 0.990 | *Dialister* | 1.08 (0.94-1.24) | 0.259 | 0.871 |
| *Lichenihabitans* | 1.07 (0.95-1.21) | 0.281 | 0.990 | *Dolosicoccus* | 1.08 (0.94-1.25) | 0.261 | 0.871 |
| *Methyloterricola* | 0.93 (0.80-1.07) | 0.317 | 0.990 | *F0540* | 0.92 (0.79-1.07) | 0.269 | 0.871 |
| *Brochothrix* | 0.88 (0.68-1.13) | 0.317 | 0.990 | *Brochothrix* | 0.86 (0.65-1.14) | 0.289 | 0.871 |
| *Sulfuricurvum* | 0.93 (0.79-1.08) | 0.329 | 0.990 | *Lichenihabitans* | 1.07 (0.94-1.22) | 0.303 | 0.871 |
| *VFKE01* | 0.88 (0.67-1.16) | 0.355 | 0.990 | *Cloacibacterium* | 0.93 (0.79-1.08) | 0.324 | 0.871 |
| *Dolosicoccus* | 1.07 (0.92-1.23) | 0.378 | 0.990 | *Methyloterricola* | 0.92 (0.77-1.09) | 0.335 | 0.871 |
| *Dialister* | 1.06 (0.92-1.22) | 0.406 | 0.990 | *Sulfuricurvum* | 0.91 (0.75-1.11) | 0.344 | 0.871 |
| *Bruticola* | 0.94 (0.80-1.10) | 0.429 | 0.990 | *Bruticola* | 0.93 (0.78-1.10) | 0.380 | 0.902 |
| *F0540* | 0.95 (0.84-1.08) | 0.455 | 0.990 | *DSSC01* | 0.92 (0.75-1.12) | 0.415 | 0.928 |
| *DSSC01* | 0.94 (0.78-1.12) | 0.487 | 0.990 | *Methanofastidiosum* | 0.93 (0.76-1.14) | 0.499 | 0.990 |
| *Oliverpabstia* | 1.03 (0.95-1.12) | 0.519 | 0.990 | *Neobacillus* | 0.96 (0.83-1.11) | 0.554 | 0.990 |
| *Methanofastidiosum* | 0.95 (0.80-1.13) | 0.544 | 0.990 | *Desulfofarcimen* | 0.95 (0.80-1.14) | 0.584 | 0.990 |
| *Cloacibacterium* | 0.96 (0.83-1.11) | 0.559 | 0.990 | *Oliverpabstia* | 1.02 (0.94-1.12) | 0.591 | 0.990 |

| | | | | | | | |
|---|---|---|---|---|---|---|---|
| *Roseburia* | 1.03 (0.91-1.17) | 0.604 | 0.990 | *Psychrosphaera* | 0.96 (0.81-1.13) | 0.592 | 0.990 |
| *UPXZ01* | 1.03 (0.88-1.22) | 0.695 | 0.990 | *RUG13038* | 1.03 (0.89-1.19) | 0.734 | 0.990 |
| *Bisgaardia* | 1.01 (0.92-1.12) | 0.765 | 0.990 | *UBA4951* | 1.02 (0.90-1.15) | 0.759 | 0.990 |
| *UBA4951* | 1.02 (0.90-1.16) | 0.766 | 0.990 | *Parasutterella* | 0.98 (0.87-1.11) | 0.786 | 0.990 |
| *RUG13038* | 1.02 (0.87-1.20) | 0.776 | 0.990 | *Proteus* | 1.02 (0.90-1.14) | 0.787 | 0.990 |
| *Neobacillus* | 0.98 (0.84-1.14) | 0.781 | 0.990 | *UBA3789* | 0.98 (0.85-1.14) | 0.792 | 0.990 |
| *Desulfofarcimen* | 0.98 (0.83-1.15) | 0.782 | 0.990 | *Bisgaardia* | 1.01 (0.92-1.12) | 0.794 | 0.990 |
| *Psychrosphaera* | 0.98 (0.86-1.12) | 0.785 | 0.990 | *Acetatifactor* | 1.02 (0.90-1.15) | 0.800 | 0.990 |
| *Olegusella* | 1.02 (0.87-1.19) | 0.817 | 0.990 | *Salinivirga* | 0.98 (0.85-1.13) | 0.806 | 0.990 |
| *Schleiferilactobacillus* | 1.02 (0.89-1.16) | 0.826 | 0.990 | *Schleiferilactobacillus* | 1.01 (0.90-1.14) | 0.845 | 0.990 |
| *JAAAOM01* | 1.01 (0.89-1.15) | 0.889 | 0.990 | *DUUS01* | 1.01 (0.87-1.18) | 0.868 | 0.990 |
| *Parasutterella* | 1.00 (0.88-1.14) | 0.941 | 0.990 | *VFKE01* | 0.98 (0.82-1.18) | 0.871 | 0.990 |
| *Bosea* | 1.00 (0.85-1.16) | 0.955 | 0.990 | *Bosea* | 1.01 (0.87-1.17) | 0.904 | 0.990 |
| *Proteus* | 1.00 (0.89-1.12) | 0.967 | 0.990 | *JAAAOM01* | 0.99 (0.87-1.14) | 0.920 | 0.990 |
| *DUUS01* | 1.00 (0.84-1.19) | 0.975 | 0.990 | *UPXZ01* | 1.00 (0.85-1.17) | 0.972 | 0.990 |
| *Salinivirga* | 1.00 (0.87-1.15) | 0.983 | 0.990 | *Olegusella* | 1.00 (0.87-1.16) | 0.981 | 0.990 |
| *UBA3789* | 1.00 (0.85-1.17) | 0.990 | 0.990 | *Roseburia* | 1.00 (0.88-1.13) | 0.990 | 0.990 |

Results are based on the Fine-Gray model adjusted for Sex, APOE genotype (0 vs 1-2 ε alleles), Education level, BMI, Systolic blood pressure, Alcohol consumption, Smoking status, Usage of Anti-Hypertensive Medication, Gut microbiome altering drugs, Prevalent Diabetes, Prevalent CVD. Age was used as the time scale.

Supplementary Table 11. Associations between species linked to APOE genotype and Alzheimer's disease or dementia. The results are ordered based on p-value.

| | Alzheimer's Disease | | | | Dementia | | |
|---|---|---|---|---|---|---|---|
| Predictor | HR (95%CI) | P-value | q-value | Predictor | HR (95%CI) | P-value | q-value |
| *Nocardia carnea* | 1.08 (1.03-1.14) | 0.002 | 0.172 | *Nocardia carnea* | 1.13 (1.07-1.19) | <0.001 | <0.001 |
| *Akkermansia glycaniphila* | 1.21 (1.05-1.40) | 0.009 | 0.315 | *Akkermansia glycaniphila* | 1.21 (1.05-1.39) | 0.007 | 0.301 |
| *JAAYUG01 sp012517825* | 0.76 (0.62-0.94) | 0.011 | 0.315 | *JAAYUG01 sp012517825* | 0.76 (0.61-0.95) | 0.016 | 0.459 |
| *Propionibacterium cyclohexanicum* | 0.87 (0.76-0.99) | 0.032 | 0.452 | *Ectobacillus funiculus* | 0.85 (0.73-0.99) | 0.040 | 0.722 |
| *Ectobacillus funiculus* | 0.87 (0.77-0.99) | 0.033 | 0.452 | *Propionibacterium cyclohexanicum* | 0.84 (0.71-0.99) | 0.042 | 0.722 |
| *Roseburia inulinivorans* | 1.18 (1.01-1.37) | 0.035 | 0.452 | *Marinococcus luteus* | 0.84 (0.69-1.02) | 0.073 | 0.778 |
| *Pedobacter insulae* | 1.11 (1.01-1.22) | 0.037 | 0.452 | *UBA3947 sp900095445* | 1.12 (0.99-1.28) | 0.077 | 0.778 |
| *Marinococcus luteus* | 0.80 (0.65-0.99) | 0.042 | 0.452 | *Pedobacter insulae* | 1.10 (0.99-1.24) | 0.087 | 0.778 |
| *Winkia sp002849225* | 0.75 (0.54-1.04) | 0.086 | 0.822 | *Lentilactobacillus hilgardii* | 1.11 (0.99-1.24) | 0.087 | 0.778 |
| *Lactobacillus melliventris* | 1.08 (0.98-1.20) | 0.109 | 0.839 | *Advenella mimigardefordensis* | 1.08 (0.98-1.17) | 0.107 | 0.778 |
| *Salimicrobium jeotgali* | 0.90 (0.78-1.03) | 0.124 | 0.839 | *Winkia sp002849225* | 0.78 (0.58-1.06) | 0.110 | 0.778 |
| *Fundidesulfovibrio putealis* | 0.83 (0.66-1.05) | 0.125 | 0.839 | *Prevotella sp000688375* | 1.12 (0.97-1.30) | 0.132 | 0.778 |
| *Prevotella sp000688375* | 1.13 (0.96-1.32) | 0.131 | 0.839 | *Roseburia inulinivorans* | 1.12 (0.97-1.30) | 0.134 | 0.778 |
| *Neobacillus thermocopriae* | 0.91 (0.79-1.05) | 0.191 | 0.839 | *Ralstonia pseudosolanacearum* | 1.09 (0.97-1.23) | 0.146 | 0.778 |
| *Acetatifactor sp900066565* | 1.09 (0.96-1.23) | 0.193 | 0.839 | *Fundidesulfovibrio putealis* | 0.89 (0.75-1.04) | 0.149 | 0.778 |
| *Advenella mimigardefordensis* | 0.92 (0.82-1.04) | 0.194 | 0.839 | *Salimicrobium jeotgali* | 0.88 (0.73-1.05) | 0.155 | 0.778 |
| *Lentilactobacillus hilgardii* | 1.10 (0.95-1.28) | 0.206 | 0.839 | *Arcticibacter svalbardensis* | 0.90 (0.77-1.05) | 0.177 | 0.778 |
| *Prevotella oris* | 1.10 (0.95-1.27) | 0.212 | 0.839 | *Lactobacillus melliventris* | 1.09 (0.95-1.24) | 0.210 | 0.778 |
| *Ralstonia pseudosolanacearum* | 1.06 (0.96-1.18) | 0.216 | 0.839 | *Rothia aeria* | 0.92 (0.80-1.05) | 0.212 | 0.778 |
| *Brevibacterium senegalense* | 0.90 (0.76-1.07) | 0.230 | 0.839 | *Neobacillus thermocopriae* | 0.90 (0.76-1.07) | 0.219 | 0.778 |

| | | | | | | | |
|---|---|---|---|---|---|---|---|
| *Dermacoccus profundi* | 0.92 (0.80-1.06) | 0.233 | 0.839 | *Salinivibrio socompensis* | 1.05 (0.97-1.12) | 0.225 | 0.778 |
| *UBA3947 sp900095445* | 1.08 (0.95-1.23) | 0.239 | 0.839 | *Prevotella micans* | 1.09 (0.95-1.24) | 0.231 | 0.778 |
| *Prevotella micans* | 1.09 (0.94-1.26) | 0.252 | 0.839 | *Staphylococcus capitis* | 0.89 (0.72-1.08) | 0.238 | 0.778 |
| *Arcticibacter svalbardensis* | 0.92 (0.80-1.06) | 0.255 | 0.839 | *Aeromicrobium choanae* | 1.06 (0.96-1.16) | 0.250 | 0.778 |
| *Rothia aeria* | 0.92 (0.79-1.07) | 0.258 | 0.839 | *UBA3789 sp002369415* | 0.91 (0.78-1.07) | 0.251 | 0.778 |
| *Ilyobacter polytropus* | 0.89 (0.72-1.10) | 0.265 | 0.839 | *RUG13038 sp902789425* | 1.09 (0.94-1.25) | 0.254 | 0.778 |
| *Ligilactobacillus apodemi* | 0.92 (0.79-1.08) | 0.300 | 0.839 | *Dolosicoccus paucivorans* | 1.08 (0.94-1.25) | 0.254 | 0.778 |
| *Staphylococcus capitis* | 0.91 (0.76-1.10) | 0.323 | 0.839 | *Dermacoccus profundi* | 0.91 (0.76-1.07) | 0.259 | 0.778 |
| *Actinomyces slackii* | 0.92 (0.79-1.08) | 0.323 | 0.839 | *Prevotella oris* | 1.08 (0.94-1.24) | 0.270 | 0.778 |
| *Methyloterricola oryzae* | 0.93 (0.80-1.08) | 0.325 | 0.839 | *F0540 sp000466585* | 0.92 (0.79-1.07) | 0.280 | 0.778 |
| *Brochothrix thermosphacta* | 0.88 (0.68-1.13) | 0.326 | 0.839 | *Brochothrix thermosphacta* | 0.86 (0.65-1.14) | 0.299 | 0.778 |
| *Sulfuricurvum sp002327465* | 0.93 (0.79-1.08) | 0.339 | 0.839 | *Lactiplantibacillus xiangfangensis* | 1.07 (0.94-1.21) | 0.301 | 0.778 |
| *Brochothrix campestris* | 0.93 (0.81-1.08) | 0.359 | 0.839 | *Dysgonomonas capnocytophagoides* | 0.91 (0.77-1.09) | 0.303 | 0.778 |
| *Fimenecus sp900315985* | 0.94 (0.81-1.08) | 0.365 | 0.839 | *Paracoccus yeei* | 0.90 (0.74-1.10) | 0.310 | 0.778 |
| *Dolosicoccus paucivorans* | 1.07 (0.93-1.23) | 0.365 | 0.839 | *Lacticaseibacillus pantheris* | 1.05 (0.95-1.17) | 0.319 | 0.778 |
| *VFKE01 sp009885695* | 0.88 (0.66-1.16) | 0.368 | 0.839 | *Cloacibacterium normanense* | 0.93 (0.79-1.08) | 0.330 | 0.778 |
| *Corynebacterium simulans* | 0.93 (0.78-1.10) | 0.377 | 0.839 | *Methyloterricola oryzae* | 0.92 (0.76-1.10) | 0.341 | 0.778 |
| *Pediococcus inopinatus* | 0.93 (0.79-1.09) | 0.379 | 0.839 | *Sulfuricurvum sp002327465* | 0.90 (0.73-1.12) | 0.352 | 0.778 |
| *RUG13038 sp902789425* | 1.07 (0.91-1.27) | 0.407 | 0.839 | *Vibrio sp001506075* | 1.02 (0.97-1.08) | 0.365 | 0.778 |
| *CAG 313 sp000433035* | 1.07 (0.91-1.25) | 0.411 | 0.839 | *Brochothrix campestris* | 0.92 (0.76-1.11) | 0.370 | 0.778 |
| *Lactiplantibacillus xiangfangensis* | 1.06 (0.92-1.23) | 0.412 | 0.839 | *Fimenecus sp900315985* | 0.94 (0.82-1.08) | 0.377 | 0.778 |
| *UBA3789 sp002369415* | 0.93 (0.79-1.10) | 0.416 | 0.839 | *Enteromonas sp002472275* | 1.07 (0.92-1.23) | 0.381 | 0.778 |
| *Paracoccus yeei* | 0.92 (0.76-1.12) | 0.422 | 0.839 | *Ilyobacter polytropus* | 0.92 (0.77-1.11) | 0.395 | 0.778 |
| *Ligilactobacillus agilis* | 0.94 (0.82-1.09) | 0.429 | 0.839 | *Bruticola papionis* | 0.93 (0.78-1.11) | 0.398 | 0.778 |

| | | | | | | | |
|---|---|---|---|---|---|---|---|
| *Bruticola papionis* | 0.94 (0.80-1.10) | 0.448 | 0.846 | *Ligilactobacillus apodemi* | 0.94 (0.81-1.09) | 0.424 | 0.810 |
| *FEB 22 sp003105195* | 1.02 (0.96-1.09) | 0.453 | 0.846 | *DSSC01 sp011331305* | 0.92 (0.74-1.14) | 0.444 | 0.830 |
| *Ligilactobacillus murinus* | 0.95 (0.82-1.09) | 0.464 | 0.846 | *Ligilactobacillus equi* | 1.05 (0.92-1.21) | 0.462 | 0.845 |
| *F0540 sp000466585* | 0.96 (0.84-1.08) | 0.472 | 0.846 | *Pediococcus inopinatus* | 0.95 (0.83-1.09) | 0.480 | 0.851 |
| *Oliverpabstia intestinalis* | 1.03 (0.95-1.12) | 0.507 | 0.877 | *Methanofastidiosum sp001587595* | 0.93 (0.76-1.14) | 0.506 | 0.851 |
| *Dysgonomonas capnocytophagoides* | 0.95 (0.83-1.10) | 0.510 | 0.877 | *Ligilactobacillus agilis* | 0.96 (0.83-1.10) | 0.517 | 0.851 |
| *DSSC01 sp011331305* | 0.94 (0.78-1.13) | 0.526 | 0.883 | *Liquorilactobacillus uvarum* | 1.05 (0.91-1.20) | 0.525 | 0.851 |
| *Alloscardovia macacae* | 0.94 (0.78-1.14) | 0.534 | 0.883 | *Actinomyces slackii* | 0.96 (0.82-1.11) | 0.568 | 0.851 |
| *Methanofastidiosum sp001587595* | 0.95 (0.80-1.13) | 0.554 | 0.899 | *Flavobacterium cauense* | 0.94 (0.76-1.16) | 0.573 | 0.851 |
| *Cloacibacterium normanense* | 0.96 (0.83-1.11) | 0.568 | 0.905 | *CAG 313 sp000433035* | 1.04 (0.90-1.21) | 0.578 | 0.851 |
| *Bifidobacterium psychraerophilum* | 0.96 (0.82-1.12) | 0.599 | 0.913 | *Oliverpabstia intestinalis* | 1.03 (0.94-1.12) | 0.582 | 0.851 |
| *Bifidobacterium swidsinskii* | 0.96 (0.83-1.12) | 0.609 | 0.913 | *UBA4951 sp900542595* | 0.96 (0.84-1.10) | 0.592 | 0.851 |
| *Enteromonas sp002472275* | 1.04 (0.89-1.21) | 0.614 | 0.913 | *Desulfofarcimen acetoxidans* | 0.95 (0.80-1.14) | 0.599 | 0.851 |
| *Aeromicrobium choanae* | 1.02 (0.94-1.11) | 0.626 | 0.913 | *FEB 22 sp003105195* | 1.02 (0.94-1.12) | 0.607 | 0.851 |
| *Pedobacter sp001027745* | 1.02 (0.93-1.12) | 0.641 | 0.913 | *Parasutterella excrementihominis* | 0.97 (0.85-1.10) | 0.608 | 0.851 |
| *Vibrio sp001506075* | 1.01 (0.96-1.07) | 0.655 | 0.913 | *Psychrosphaera saromensis* | 0.96 (0.82-1.13) | 0.609 | 0.851 |
| *Schleiferilactobacillus perolens* | 0.97 (0.84-1.12) | 0.655 | 0.913 | *Schleiferilactobacillus perolens* | 0.97 (0.85-1.10) | 0.622 | 0.851 |
| *UBA4951 sp900542595* | 0.97 (0.85-1.11) | 0.667 | 0.913 | *Ligilactobacillus murinus* | 0.97 (0.84-1.11) | 0.623 | 0.851 |
| *Flavobacterium cauense* | 0.96 (0.81-1.15) | 0.669 | 0.913 | *Bifidobacterium swidsinskii* | 0.97 (0.84-1.11) | 0.627 | 0.851 |
| *Ligilactobacillus animalis* | 0.97 (0.84-1.12) | 0.702 | 0.943 | *Bifidobacterium crudilactis* | 1.04 (0.89-1.20) | 0.633 | 0.851 |
| *Liquorilactobacillus uvarum* | 1.03 (0.88-1.20) | 0.716 | 0.945 | *Brevibacterium senegalense* | 1.05 (0.85-1.29) | 0.645 | 0.853 |
| *Ligilactobacillus equi* | 1.03 (0.89-1.18) | 0.728 | 0.945 | *Alloscardovia macacae* | 0.96 (0.80-1.15) | 0.665 | 0.867 |
| *Bifidobacterium crudilactis* | 0.97 (0.83-1.14) | 0.741 | 0.945 | *Corynebacterium simulans* | 0.97 (0.82-1.14) | 0.700 | 0.895 |
| *Bisgaardia hudsonensis* | 1.02 (0.92-1.12) | 0.747 | 0.945 | *UPXZ01 sp900498215* | 0.97 (0.83-1.14) | 0.708 | 0.895 |

| | | | | | | | |
|---|---|---|---|---|---|---|---|
| *Parasutterella excrementihominis* | 0.98 (0.86-1.12) | 0.797 | 0.968 | *Acetatifactor sp900066565* | 1.02 (0.90-1.15) | 0.741 | 0.895 |
| *Desulfofarcimen acetoxidans* | 0.98 (0.84-1.15) | 0.801 | 0.968 | *Pediococcus stilesii* | 1.03 (0.87-1.21) | 0.744 | 0.895 |
| *Olegusella massiliensis* | 1.02 (0.87-1.19) | 0.806 | 0.968 | *Bifidobacterium psychraerophilum* | 0.98 (0.84-1.14) | 0.769 | 0.895 |
| *Psychrosphaera saromensis* | 0.98 (0.86-1.12) | 0.811 | 0.968 | *Lentilactobacillus farraginis* | 1.02 (0.88-1.19) | 0.776 | 0.895 |
| *Dialister invisus* | 0.98 (0.85-1.14) | 0.837 | 0.968 | *Bisgaardia hudsonensis* | 1.01 (0.92-1.12) | 0.780 | 0.895 |
| *Lacticaseibacillus pantheris* | 0.99 (0.89-1.10) | 0.855 | 0.968 | *Aggregatibacter actinomycetemcomitans* | 0.98 (0.87-1.11) | 0.800 | 0.895 |
| *UBA3789 sp902780585* | 1.01 (0.87-1.19) | 0.857 | 0.968 | *Pedobacter sp001027745* | 1.02 (0.90-1.15) | 0.802 | 0.895 |
| *Cellulosilyticum ruminicola* | 1.01 (0.87-1.18) | 0.867 | 0.968 | *Proteus mirabilis* | 1.01 (0.90-1.14) | 0.806 | 0.895 |
| *JAAAOM01 sp009908855* | 1.01 (0.89-1.15) | 0.869 | 0.968 | *Salinivirga cyanobacteriivorans* | 0.98 (0.85-1.13) | 0.811 | 0.895 |
| *Lentilactobacillus farraginis* | 1.01 (0.85-1.21) | 0.878 | 0.968 | *Dwaynesavagella sp000283555* | 0.99 (0.88-1.10) | 0.812 | 0.895 |
| *Pediococcus stilesii* | 1.01 (0.85-1.19) | 0.935 | 0.989 | *Ligilactobacillus animalis* | 0.98 (0.85-1.14) | 0.825 | 0.898 |
| *Proteus mirabilis* | 1.00 (0.89-1.12) | 0.936 | 0.989 | *DUUS01 sp012841465* | 1.01 (0.87-1.18) | 0.863 | 0.928 |
| *Salinivirga cyanobacteriivorans* | 1.00 (0.87-1.15) | 0.976 | 0.989 | *VFKE01 sp009885695* | 0.99 (0.82-1.18) | 0.888 | 0.943 |
| *UPXZ01 sp900498215* | 1.00 (0.85-1.19) | 0.981 | 0.989 | *Cellulosilyticum ruminicola* | 0.99 (0.86-1.15) | 0.901 | 0.945 |
| *DUUS01 sp012841465* | 1.00 (0.84-1.19) | 0.984 | 0.989 | *JAAAOM01 sp009908855* | 0.99 (0.87-1.14) | 0.933 | 0.965 |
| *Dwaynesavagella sp000283555* | 1.00 (0.91-1.09) | 0.984 | 0.989 | *UBA3789 sp902780585* | 0.99 (0.86-1.15) | 0.943 | 0.965 |
| *Salinivibrio socompensis* | 1.00 (0.91-1.10) | 0.987 | 0.989 | *Dialister invisus* | 1.00 (0.87-1.15) | 0.963 | 0.974 |
| *Aggregatibacter actinomycetemcomitans* | 1.00 (0.89-1.13) | 0.989 | 0.989 | *Olegusella massiliensis* | 1.00 (0.87-1.16) | 0.989 | 0.989 |

Results are based on the Fine-Gray model adjusted for Sex, APOE genotype (0 vs 1-2 ε alleles), Education level, BMI, Systolic blood pressure, Alcohol consumption, Smoking status, Usage of Anti-Hypertensive Medication, Gut microbiome altering drugs, Prevalent Diabetes, Prevalent CVD. Age was used as the time scale.

Supplementary Table 12. Performance metrics of the XGBoost model, reported as mean (SD).

Dementia

| | Accuracy | Sensitivity | Specificity | AUC |
|---|---|---|---|---|
| Clinival covariates only | 0.920 (0.005) | 0.062 (0.019) | 0.996 (0.002) | 0.840 (0.014) |
| Phylum | 0.919 (0.005) | 0.052 (0.017) | 0.997 (0.002) | 0.835 (0.014) |
| Genus | 0.918 (0.005) | 0.031 (0.018) | 0.997 (0.002) | 0.829 (0.014) |
| Species | 0.919 (0.005) | 0.034 (0.018) | 0.997 (0.002) | 0.827 (0.014) |

Alzheimer's disease

| | Accuracy | Sensitivity | Specificity | AUC |
|---|---|---|---|---|
| Clinival covariates only | 0.932 (0.005) | 0.064 (0.019) | 0.997 (0.002) | 0.843 (0.014) |
| Phylum | 0.932 (0.005) | 0.049 (0.019) | 0.997 (0.002) | 0.837 (0.014) |
| Genus | 0.931 (0.005) | 0.028 (0.018) | 0.998 (0.001) | 0.832 (0.013) |
| Species | 0.930 (0.005) | 0.028 (0.017) | 0.998 (0.001) | 0.833 (0.013) |

Supplementary Table 13. XGBoost-identified contributing factors in descending order.

Dementia

| Clinival covariates only | Phylum | Genus | Species |
|---|---|---|---|
| Age (0.438) | Age (0.247) | Age (0.129) | Age (0.120) |
| APOE genotype (0.188) | APOE genotype (0.140) | APOE genotype (0.062) | APOE genotype (0.056) |
| Alcohol consumption (0.085) | Alcohol consumption (0.056) | *Granulicoccus* (0.024) | *Prevotella oulorum* (0.016) |
| Systolic blood pressure (0.082) | Systolic blood pressure (0.055) | *UBA6382* (0.023) | *Granulicoccus phenolivorans* (0.016) |
| Gut microbiome altering drugs (0.056) | Gut microbiome altering drugs (0.037) | Systolic blood pressure (0.021) | *Bifidobacterium animalis* (0.014) |
| BMI (0.047) | *Bacteroidota* (0.031) | *Lactococcus A 346120* (0.016) | *Bacteroides H salyersiae* (0.013) |
| Anti-Hypertensive Medication (0.044) | *Patescibacteria* (0.030) | Alcohol consumption (ALKI2) (0.012) | *Bacteroides H stercoris* (0.013) |
| Education level (0.024) | *Desulfobacterota I* (0.029) | *Aphodousia* (0.012) | *Bacteroides H xylanisolvens* (0.012) |
| Smoking status (0.019) | BMI (0.028) | *Ligilactobacillus* (0.011) | *Parabacteroides B 862066 goldsteinii* (0.012) |
| Prevalent CVD (0.008) | *Planctomycetota* (0.026) | D*16.34* (0.011) | *Pseudomonas E 650326 knackmussii* (0.011) |
| Sex (0.005) | *Actinobacteriota* (0.025) | *F0058* (0.010) | *Lactococcus A 346120 lactis 344179* (0.010) |
| Antibiotics used in the baseline (0.003) | Anti-Hypertensive Medication (0.024) | *Pseudomonas F* (0.010) | *Aphodousia faecalis* (0.010) |
| Prevalent diabetes (0.002) | *Proteobacteria* (0.023) | *Paratractidigestivibacter* (0.009) | *F0058 sp000768855* (0.009) |
| | *Fusobacteriota* (0.023) | *Lactobacillus* (0.009) | *Prevotella multisaccharivorax* (0.009) |
| | *Cyanobacteria* (0.021) | *SFLA01* (0.009) | *Dielma fastidiosa* (0.008) |
| | *Firmicutes C* (0.020) | *CAG 710* (0.009) | Liquorilactobacillus vini (0.007) |
| | *Firmicutes A* (0.016) | *Faecenecus* (0.008) | *Limosilactobacillus timonensis* (0.007) |
| | *Synergistota* (0.016) | *Aphodomorpha* (0.008) | Systolic blood pressure (0.007) |
| | *Verrucomicrobiota* (0.013) | *WSTA01* (0.008) | Alcohol consumption (ALKI2) (0.007) |
| | *Campylobacterota* (0.012) | *Vaginella* (0.008) | *Faecenecus gallistercoris* (0.007) |
| | *Firmicutes B 370514* (0.011) | *Parabacteroides B 862066* (0.008) | *Limosilactobacillus vaginalis* (0.007) |
| | *Fibrobacterota* (0.011) | *CAG 267* (0.007) | *Phocaeicola A 858004 salanitronis* (0.007) |
| | *Methanobacteriota A 1229* (0.010) | *CAG 41* (0.007) | *D16 34 sp 009911635* (0.006) |
| | *Firmicutes D* (0.010) | *Parascardovia* (0.007) | |

| Alzheimer's disease | | | |
|---|---|---|---|
| Clinival covariates only | Phylum | Genus | Species |
| Age (0.425) | Age (0.240) | Age (0.118) | Age (0.110) |
| APOE genotype (0.226) | APOE genotype (0.163) | APOE genotype (0.069) | APOE genotype (0.062) |
| Alcohol consumption (ALKI2) (0.073) | Alcohol consumption (0.043) | *Granulicoccus* (0.026) | *Parabacteroides B 862066 goldsteinii* (0.023) |
| Systolic blood pressure (0.066) | Patescibacteria (0.037) | *Pseudomonas F* (0.020) | *Bacteroides H salyersiae* (0.023) |
| BMI (0.050) | Systolic blood pressure (0.035) | *Aphodousia* (0.018) | *Prevotella oulorum* (0.017) |
| Education level (0.037) | *Bacteroidota* (0.030) | *UBA6382* (0.017) | *Granulicoccus phenolivorans* (0.016) |
| Gut microbiome altering drugs (0.033) | *Firmicutes C* (0.030) | *Lactococcus A 346120* (0.016) | *Aphodousia faecalis* (0.016) |
| Anti-Hypertensive Medication (0.032) | *Desulfobacterota I* (0.028) | *Faecenecus* (0.015) | *Prevotella multisaccharivorax* (0.015) |
| Smoking status (0.025) | *Cyanobacteria* (0.025) | *Parascardovia* (0.015) | *Pseudomonas F resinovorans_A* (0.014) |
| Prevalent CVD (0.014) | *Actinobacteriota* (0.023) | *ER4* (0.013) | *Faecenecus gallistercoris* (0.012) |
| Sex (0.013) | *Synergistota* (0.023) | *SFLA01* (0.013) | *SFLA01 sp004553575* (0.010) |
| Prevalent diabetes (0.004) | *Proteobacteria* (0.022) | *Ligilactobacillus* (0.011) | *Limosilactobacillus timonensis* (0.009) |
| Antibiotics used in the baseline (0.002) | BMI (0.021) | *WSTA01* (0.010) | *Lactococcus A 346120 lactis 344179* (0.009) |
| | *Fusobacteriota* (0.018) | *D16.34* (0.010) | *D16 34 sp009911635* (0.008) |
| | *Verrucomicrobiota* (0.018) | *Dialister* (0.009) | *Cryptobacteroides sp000435075* (0.008) |
| | *Chloroflexota* (0.017) | *Coprobacillus* (0.008) | *Dielma fastidiosa* (0.008) |
| | *Firmicutes A* (0.016) | *CABJAA01* (0.008) | *WSTA01 sp009789225* (0.007) |
| | Gut microbiome altering drugs (0.016) | *Aphodomorpha* (0.008) | *CABJAA01 sp003150885* (0.007) |
| | Anti-Hypertensive Medication (0.015) | *Parabacteroides_B_862066* (0.008) | *Bacteroides H fragilis* (0.006) |
| | *Planctomycetota* (0.014) | *SFTJ01* (0.008) | *Cryptobacteroides sp900544195* (0.006) |
| | Education level (0.014) | *Slackia A* (0.007) | *Liquorilactobacillus vini* (0.006) |
| | *Fibrobacterota* (0.013) | *Cryptobacteroides* (0.007) | *Ruminococcus D sp000434695* (0.006) |
| | *Firmicutes D* (0.013) | *Capnocytophaga 820688* (0.007) | *Odoribacter massiliensis* (0.006) |
| | *Methanobacteriota A 1229* (0.013) | *RuminococcusD* (0.007) | *Coprobacillus cateniformis* (0.006) |
| | *Firmicutes B 370514* (0.011) | *RZYY01* (0.007) | *Prevotella copri* (0.006) |
| | *Firmicutes B 370537* (0.011) | *CAG 273* (0.006) | *Bifidobacterium animalis* (0.005) |
| | *Spirochaetota* (0.011) | *MWCK01* (0.006) | *F0058 sp000768855* (0.005) |
| | *Firmicutes E* (0.008) | *Sodaliphilus* (0.006) | *Paraprevotella sp003477995* (0.005) |

Abbrevations: BP – Blood pressure; BMI – Body Mass Index; CVD – Cardiovascular Disease
